# Monkeypox vaccination willingness, determinants, and communication needs in gay, bisexual, and other men who have sex with men, in the context of limited vaccine availability in the Netherlands (Dutch MPX-survey)

**DOI:** 10.1101/2022.10.11.22280965

**Authors:** Nicole HTM Dukers-Muijrers, Ymke Evers, Veja Widdershoven, Udi Davidovich, Philippe CG Adam, Eline LM Op de Coul, Paul Zantkuijl, Amy Matser, Maria Prins, Henry JC de Vries, Casper den Heijer, Christian JPA Hoebe, Anne-Marie Niekamp, Francine Schneider, Juliana Reyes-Urueña, Roberto Croci, Angelo D’Ambrosio, Marc van der Valk, Dirk Posthouwer, Robin Ackens, Henriette ter Waarbeek, Teymur Noori, Elske Hoornenborg

## Abstract

**Introduction:** In the global monkeypox outbreak primary preventive vaccination is offered to people at higher risk for infection. We study vaccine acceptance and its determinants, to target and tailor public health (communication-)strategies in the context of limited vaccine supply in the Netherlands. Methods. Online survey in a convenience sample of gay, bisexual and other men who have sex with men, including transgender persons (22/07-05/09/2022, the Netherlands). We assessed determinants (sociodemographic, social environment, medical, and behavioral factors, and beliefs) for being (un)willing to accept vaccination. We used multivariable multinominal regression and logistic regression analyses, calculating adjusted odds ratios (aOR) and 95 percent confidence-intervals. An open question asked for campaigning and procedural recommendations.

**Results:** Of respondents, 81.5% (n=1,512/1,856) were willing to accept vaccination; this was 85.2% (799/938) in vaccination-eligible people (HIV-PrEP use, living with HIV, STI, or >3 partners) and 77.7% (713/918) in those non-eligible. Determinants for non-acceptance included: urbanization (rural: aOR:2.2;1.2-3.7; low-urban: aOR:2.4;1.4-3.9; versus high-urban), not knowing monkeypox-vaccinated persons (aOR:2.4;1.6-3.4), and lack of connection to gay/queer-community (aOR:2.0;1.5-2.7). Beliefs associated with acceptance were perception of higher risk/severity of monkeypox, higher protection motivation, positive outcome expectations post vaccination (effectiveness and side-effects), and perceived positive social norms regarding vaccination of their social network.

Respondents recommended more accessible communication, delivered regularly, stigma-free, sex positive and with facts on monkeypox, vaccination benefits and procedures, and explain (other) preventive options. For vaccination, it was recommended to add ‘self-registration’, provision also at non-clinic settings, discrete/anonymous options, and more inclusive strategies to reach people (e.g., those not in existing patient-registries) at high risk for monkeypox.

**Conclusion:** In the public health response to the monkeypox outbreak, key is a broad and equitable access to information, and low-threshold vaccination options for those at highest risk.

Communication should be transparent and tailored to beliefs, such as perceived risks of monkeypox, benefits of vaccination, and social norms, and should include other preventive options. Public health efforts may be strengthened in less urbanized areas and reach out to those who lack relevant social network influences.

## INTRODUCTION

Monkeypox (MPX) outbreaks have been reported in nonendemic countries since May 2022 [1-3]. As of 20 September 2022, 64,881 confirmed cases of MPX were reported worldwide, and 19,827 from 29 EU/EEA countries, including 1,221 in the Netherlands [3,4]. The overall risk for MPX is assessed as moderate for people having multiple sexual partners (including some GBMSM/TGP) and low for the broader population [3]. Most cases have been observed in gay, bisexual and other men who have sex with men and transgender persons (GBMSM/TGP) who have multiple sexual partners [1,2]. On July 23 2022, the WHO Director-General declared the escalating global MPX outbreak a Public Health Emergency of International Concern [5]. To address these outbreaks with the required urgency, countries apply preventive measures as active case finding, contact tracing, self-isolation and quarantine.

As MPX is caused by a virus similar to smallpox, smallpox vaccines are expected to prevent or reduce the severity of the MPX infection and onward transmission, though more studies are needed to demonstrate the effect of vaccination [6-8]. Smallpox vaccine development has a long history, and various countries now offer these vaccines as post-exposure vaccination (PEPV) to contacts of a MPX case and as primary preventive pre-exposure vaccination (PPV) [9]. Scarce vaccine supplies challenge an equitable global and national public health response [10]. Countries with a limited vaccine supply, including the Netherlands, have restricted PPV-access based on high risk of MPX exposure [11]. To achieve a high vaccination coverage in people with a high risk for exposure is important to control the spread of MPX. People’s willingness to accept PPV is a vital step. For the design of successful public health strategies it is key to know the relevant determinants for vaccine acceptance [12-15]. Some determinants can inform the targeting of public health efforts to subgroups that have lower PPV acceptance. Other theory-based determinants reflect the underlying beliefs for PPV acceptance and can inform the tailoring of communication-messages.

We conducted an online survey in MPX unvaccinated GBMSM/TGP, around the start of the MPX PPV-program in the Netherlands. The survey evaluates willingness to accept PPV and its determinants, and also asked persons for their recommendations in campaigning and PPV-program procedures. These insights will contribute to shaping an equitable and inclusive public health response in the context of limited vaccine supply.

## METHODS

### Setting in the Netherlands

PEPV was already available at an earlier stage and the PPV program started at 25 July 2022 [11]. The most affected large cities started, and in the course of August, PPV activities were gradually rolled out throughout the country. A total number of vaccine doses were available to invite 32,000 people by personal email or letter, based on patient-registries of public health Center for Sexual Health (CSH), HIV outpatient clinics, or general practitioners (GP). PPV eligible were GBMSM/TGP participating in (or on a waiting list for) the national pre-exposure prophylaxis program for HIV (HIV-PrEP), were living with HIV and deemed at MPX risk by the HIV-nurse, or had according to a CSH registry in the past six months an STI diagnosis (syphilis, gonorrhea, or chlamydia), was notified for STI/HIV, or had more than three sex partners [11].

### Study design and recruitment

A prospective online survey of which baseline data are here reported (cross-sectional design). Convenience sampling was used to recruit respondents via social media channels and ‘offline’ at CSH, HIV outpatient clinics, and sex-on-premises venues. Details on recruitment channels are available in Suppl. 1. Recruitment was performed from 22 July to 5 Sept 2022, which was around the early roll-out of PPV in the Netherlands. During the survey-period, vaccination was by subcutaneous route of administration.

### Participation, ethical approval, and study population

People who were 16 years or older were eligible for participation. Participation started after providing informed consent to the study. Respondents were also asked whether they consented to be approached for later follow-up. The Medical Ethical Committee of Maastricht University waived ethical approval because the data were coded and were analyzed anonymously (METC 2022-3324). People who (ever) had sex with a man were included in analyses when they also reported (i) male sex or intersex and male, non binary, genderfluid, or agender gender identity (GBMSM), or (ii) male sex and female gender identity, or female sex and male gender identity (TGP). See Fig. 1 for the flowchart of included persons in the study population.

**Figure 1.**
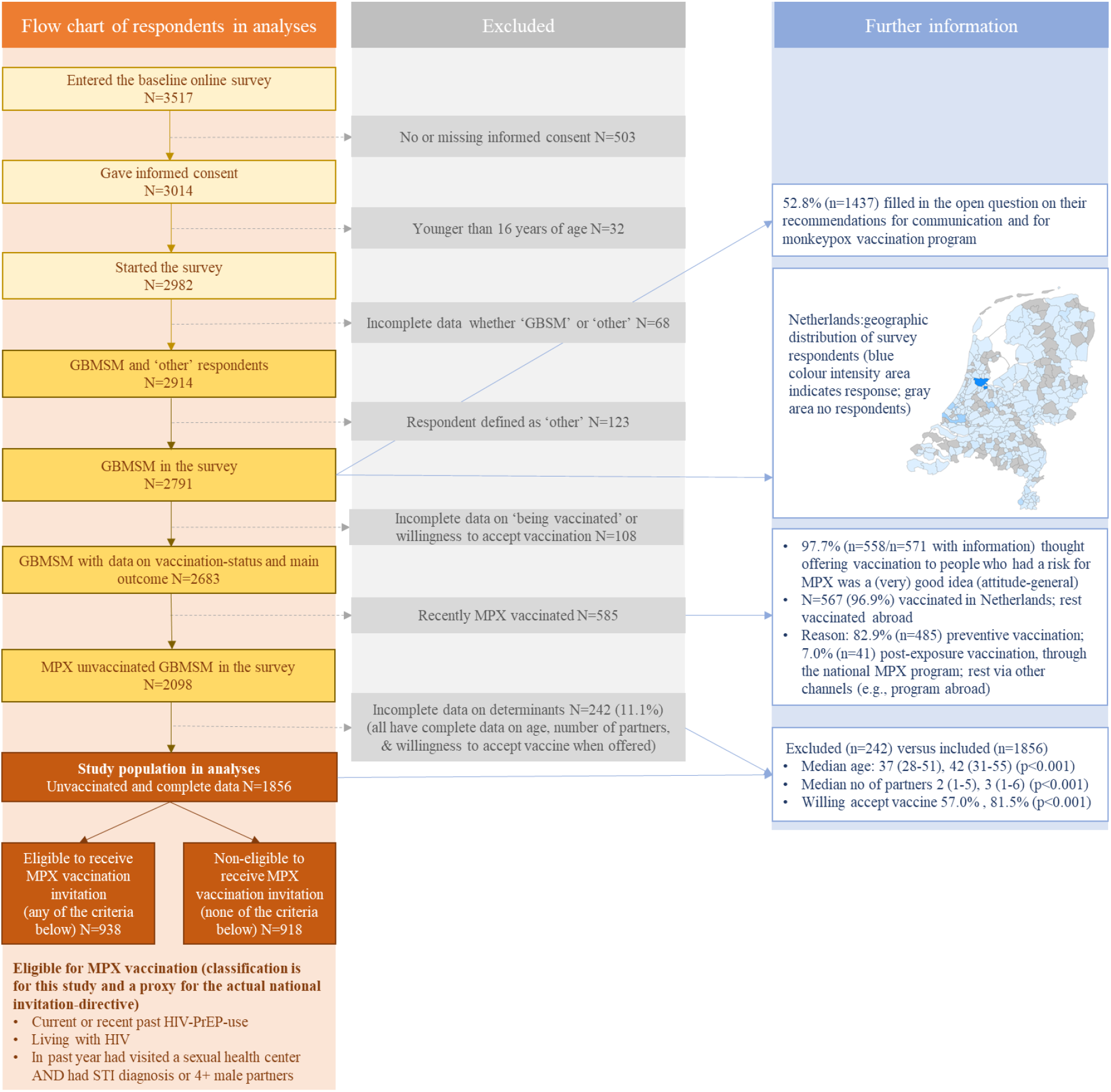
Flow chart of the number of respondents in the Dutch online MPX-survey

### Online questionnaire

The questionnaire was available in Dutch and English and its development was informed by a community consultation. Details on variables and their order (e.g., willingness was assessed early) are available in Suppl. 2.

#### Main outcome

Intention is operationalized as willingness to accept PPV when offered, by the statement ‘If you could receive a vaccine against monkeypox, would you get vaccinated against monkeypox?’ with response options 1 to 5 (Likert scale), ‘No, certainly not’, ‘No, probably not’, ‘Neutral’, ‘Yes, probably’, ‘Yes, certainly’. As a secondary outcome, willingness to accept PEPV was assessed (same Likert scale) by the statement ‘Suppose you had sex with someone with monkeypox, would you get vaccinated?’.

#### Determinants for the targeting of strategies

Sociodemographic, medical, social environment factors, and behaviors were considered as important determinants to inform the targeting of strategies to determinant-subgroups with higher PPV non-acceptance.

#### Determinants for tailoring strategies

Socio-cognitive determinants such as attitudes, cognitions and perceptions (here called: beliefs) may influence willingness to accept vaccination [16-20]. Beliefs are reputed modifiable by tailored communication-messages. Beliefs were presented in Suppl. 2, and include (i) perceived risk, severity and concern about MPX, (ii) motivation/perceived importance to protect against MPX, (iii) perceived response efficacy of vaccination, and (iv) perceived norms/social influence. Beliefs were selected based on their relevance at the time of survey-design before PPV-program start. All were theory-based derived from the Protection Motivation Theory, Health belief model, and Theory of Planned Behaviour [17-19].

#### Open question on campaigning and procedural recommendations

To collect non-guided insights in MPX communication and procedural preferences from GBMSM/TPG participants, an open question was included stating ‘What do you think is important in communication about vaccination against Monkeypox? You can, for example, indicate what and how, in your opinion, organizations can best communicate about this to people, or where you would like to get the vaccine’.

#### Eligible to receive invitation for vaccination in the Dutch PPV

Respondents were categorized as likely PPV ‘eligible’ or ‘non-eligible, which is a best proxy for the actual Dutch directive for PPV-eligibility [11]. Categorization was based on self-reported information only (not clinic registry information) and based on similar criteria as in the directive (exact criteria were not known at the time of survey-design) [11]. Respondents were categorized as ‘eligible’ when they reported (i) HIV-PrEP use in the past three months or longer ago, (ii) living with HIV (regardless of antiretroviral therapy use or sexual behavior), or (iii) visited a CSH in the past year and reported a diagnosis of chlamydia, gonorrhea, or syphilis in the past year or reported more than three male sex partners in the past three months. All other respondents not reporting any of these criteria were categorized as PPV non-eligible.

### Statistical analyses

#### Quantitatively measured data

Analyses were performed for the entire group, and for eligible and non eligible respondents. Firstly, descriptive statistics were provided on study population characteristics. Secondly, we provided descriptive statistics on the outcomes of this study, by its five categories and a priori regrouping into three categories, i.e., willing to accept PPV (certainly and likely willing), neutral, and unwilling (likely and certainly not willing). Thirdly, we evaluated non-modifiable determinants that could serve as targets of prevention strategies. Evaluated were categorical sociodemographic, medical, social environment, and behavior factors (Suppl. 2). Using univariable multinominal logistic regression analyses, the odds for being unwilling or neutral was expressed for each of the variable-categories compared to the reference category, calculating odds ratios (ORs) and 95 percent confidence intervals (CI). For multivariable analyses, we used a bidirectional stepwise procedure to identify the most important determinants. The bidirectional stepwise procedure started by a forward approach followed by backward elimination (variables with p<0.05 could stay in the model), and repeating these steps (including initial variables) until no new variables were added. This procedure was performed for the entire study population, those eligible and those non-eligible for PPV. In sensitivity analyses, we added calendar week and channel of recruitment to the multivariable models and these recruitment factors appeared not associated. Fourthly, we aimed to identify key beliefs (Suppl. 2) to inform tailoring of communication-strategies. In those eligible and in those non-eligible for PPV, we used univariable logistic regression analyses, expressing the odds of being willing to accept vaccination, for each point increase on the beliefs-scores. As aim was to assess all relevant modifiable targets to inform tailoring of communications, a separate multivariable model was constructed for each belief, adjusting for identified important non-modifiable determinants. Across analyses, we considered a P-value of <0.05 as statistically significant. All analyses were performed using SPSS package vs24 (IBM Corp., Armonk, New York, USA).

#### Open question on recommendations, qualitative data

Answers were assessed (ND, YE) and categorized into main themes that arose from the answers (inductive coding). Saturation in the answers had been reached. Comments were described per theme and illustrated with examples of citations, and where possible linked to theoretical evaluated beliefs and other determinants (deductive coding).

## RESULTS

### Respondents

Of the 2,683 GBMSM respondents, 2,098 were unvaccinated for MPX of whom 1856 (88.5%) completed the survey and 242 not completed the survey. This latter group was younger, had fewer recent sex partners, and were more often unwilling/neutral to PPV acceptance, compared to those who completed the survey (Figure 1).

### Characteristics of the study population

Of 1856 unvaccinated respondents included in analyses, 84% were born in the Netherlands, 23% lived in moderate urban to rural areas, 25% had a low/medium educational level, and median age was 42 (Tab. 1).

**Table 1.**
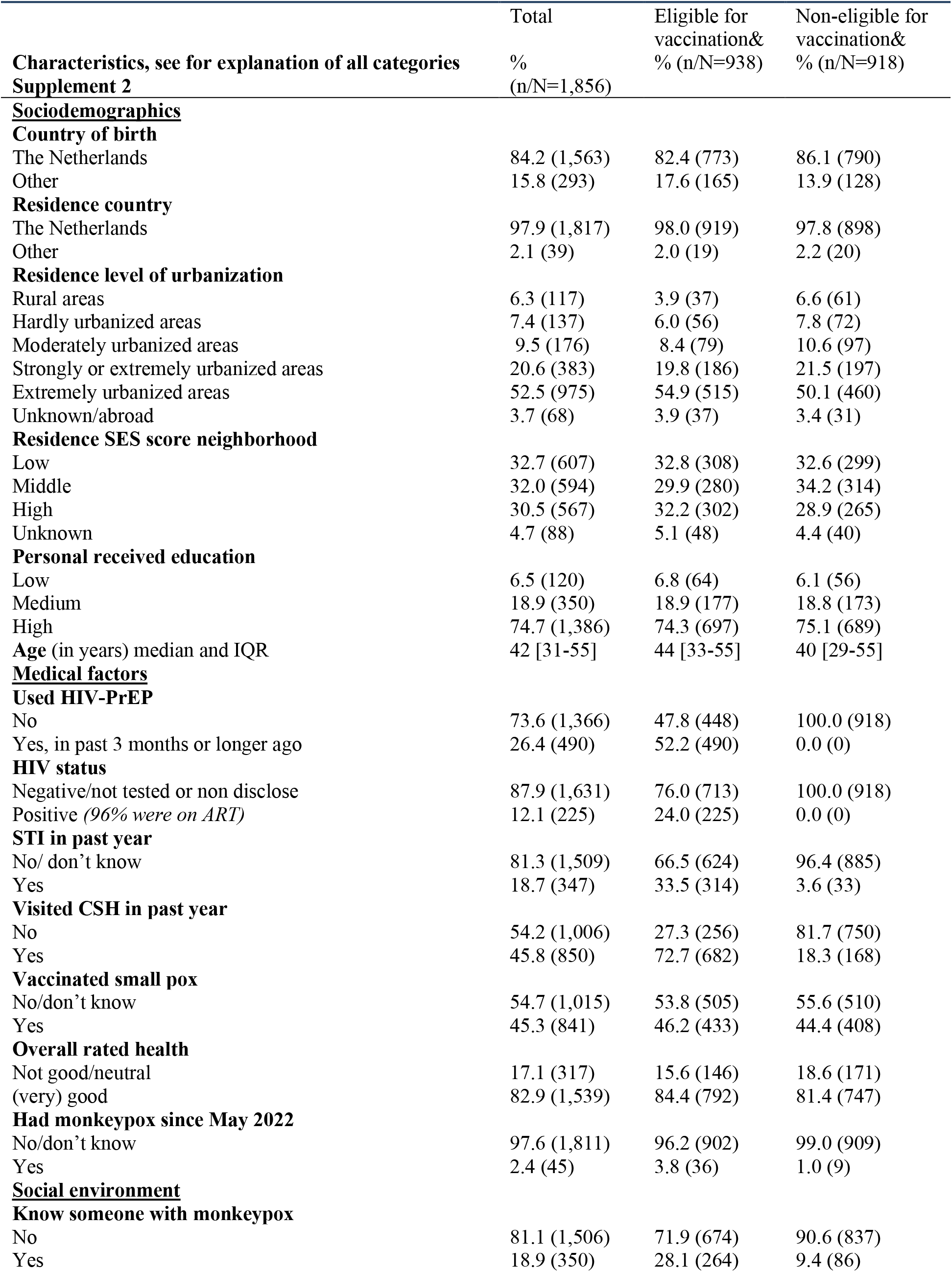
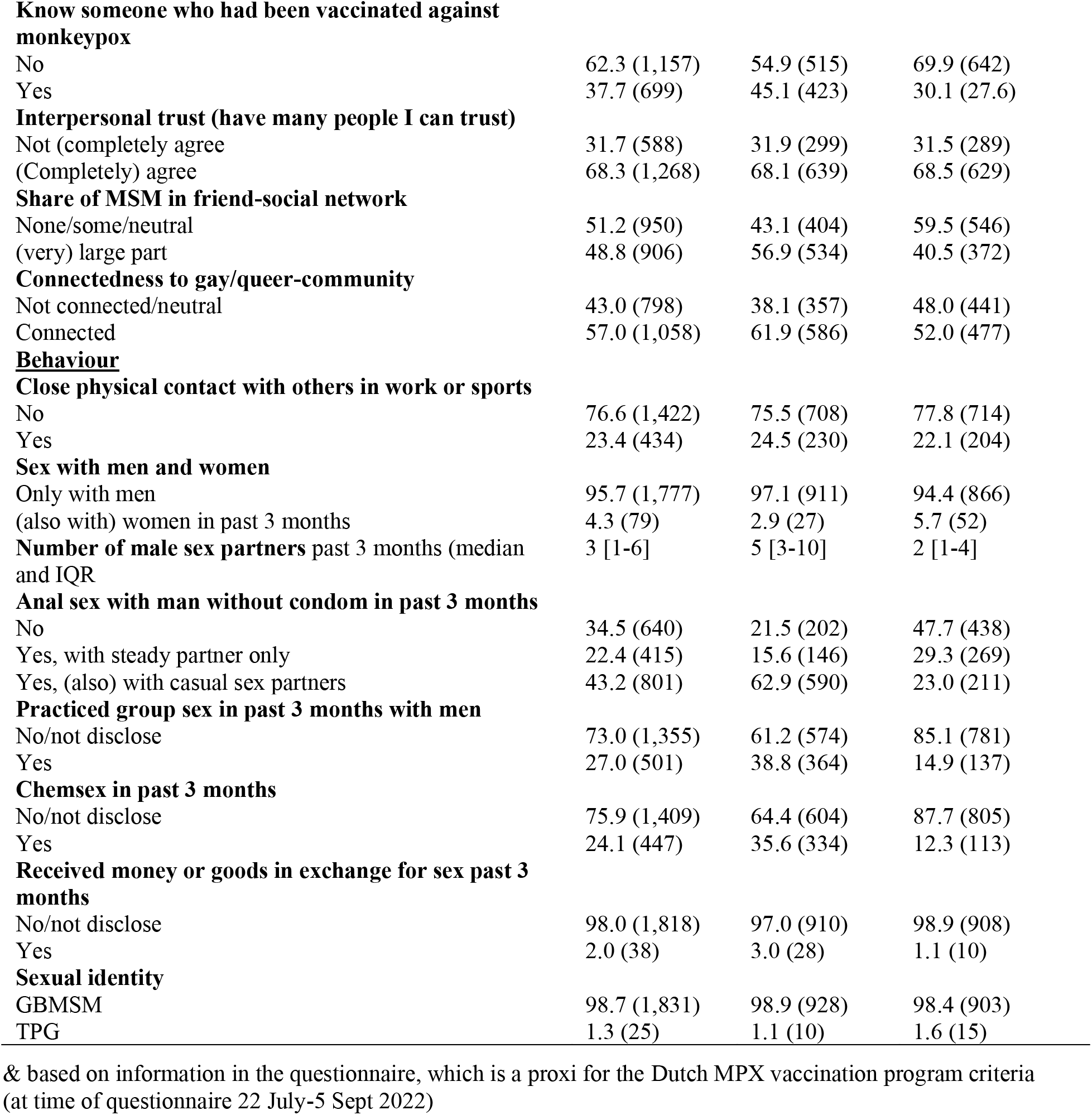
Characteristics of the study population who are unvaccinated for monkeypox; GBMSM/TGP participating in Dutch online MPX-survey

|  | Total | Eligible for vaccination& | Non-eligible for vaccination& |
| --- | --- | --- | --- |
| **Characteristics, see for explanation of all categories Supplement 2** | %<br>(n/N=1,856) | % (n/N=938) | % (n/N=918) |
| **<u>Sociodemographics</u>** |  |  |  |
| **Country of birth** |  |  |  |
| The Netherlands | 84.2 (1,563) | 82.4 (773) | 86.1 (790) |
| Other | 15.8 (293) | 17.6 (165) | 13.9 (128) |
| **Residence country** |  |  |  |
| The Netherlands | 97.9 (1,817) | 98.0 (919) | 97.8 (898) |
| Other | 2.1 (39) | 2.0 (19) | 2.2 (20) |
| **Residence level of urbanization** |  |  |  |
| Rural areas | 6.3 (117) | 3.9 (37) | 6.6 (61) |
| Hardly urbanized areas | 7.4 (137) | 6.0 (56) | 7.8 (72) |
| Moderately urbanized areas | 9.5 (176) | 8.4 (79) | 10.6 (97) |
| Strongly or extremely urbanized areas | 20.6 (383) | 19.8 (186) | 21.5 (197) |
| Extremely urbanized areas | 52.5 (975) | 54.9 (515) | 50.1 (460) |
| Unknown/abroad | 3.7 (68) | 3.9 (37) | 3.4 (31) |
| **Residence SES score neighborhood** |  |  |  |
| Low | 32.7 (607) | 32.8 (308) | 32.6 (299) |
| Middle | 32.0 (594) | 29.9 (280) | 34.2 (314) |
| High | 30.5 (567) | 32.2 (302) | 28.9 (265) |
| Unknown | 4.7 (88) | 5.1 (48) | 4.4 (40) |
| **Personal received education** |  |  |  |
| Low | 6.5 (120) | 6.8 (64) | 6.1 (56) |
| Medium | 18.9 (350) | 18.9 (177) | 18.8 (173) |
| High | 74.7 (1,386) | 74.3 (697) | 75.1 (689) |
| **Age (in years) median and IQR** | 42 [31-55] | 44 [33-55] | 40 [29-55] |
| **<u>Medical factors</u>** |  |  |  |
| **Used HIV-PrEP** |  |  |  |
| No | 73.6 (1,366) | 47.8 (448) | 100.0 (918) |
| Yes, in past 3 months or longer ago | 26.4 (490) | 52.2 (490) | 0.0 (0) |
| **HIV status** |  |  |  |
| Negative/not tested or non disclose | 87.9 (1,631) | 76.0 (713) | 100.0 (918) |
| Positive (96% were on ART) | 12.1 (225) | 24.0 (225) | 0.0 (0) |
| **STI in past year** |  |  |  |
| No/ don't know | 81.3 (1,509) | 66.5 (624) | 96.4 (885) |
| Yes | 18.7 (347) | 33.5 (314) | 3.6 (33) |
| **Visited CSH in past year** |  |  |  |
| No | 54.2 (1,006) | 27.3 (256) | 81.7 (750) |
| Yes | 45.8 (850) | 72.7 (682) | 18.3 (168) |
| **Vaccinated small pox** |  |  |  |
| No/don't know | 54.7 (1,015) | 53.8 (505) | 55.6 (510) |
| Yes | 45.3 (841) | 46.2 (433) | 44.4 (408) |
| **Overall rated health** |  |  |  |
| Not good/neutral | 17.1 (317) | 15.6 (146) | 18.6 (171) |
| (very) good | 82.9 (1,539) | 84.4 (792) | 81.4 (747) |
| **Had monkeypox since May 2022** |  |  |  |
| No/don't know | 97.6 (1,811) | 96.2 (902) | 99.0 (909) |
| Yes | 2.4 (45) | 3.8 (36) | 1.0 (9) |
| **<u>Social environment</u>** |  |  |  |
| **Know someone with monkeypox** |  |  |  |
| No | 81.1 (1,506) | 71.9 (674) | 90.6 (837) |
| Yes | 18.9 (350) | 28.1 (264) | 9.4 (86) |
| **Know someone who had been vaccinated against monkeypox** |  |  |  |
| No | 62.3 (1,157) | 54.9 (515) | 69.9 (642) |
| Yes | 37.7 (699) | 45.1 (423) | 30.1 (27.6) |
| **Interpersonal trust (have many people I can trust)** |  |  |  |
| Not (completely) agree | 31.7 (588) | 31.9 (299) | 31.5 (289) |
| (Completely) agree | 68.3 (1,268) | 68.1 (639) | 68.5 (629) |
| **Share of MSM in friend-social network** |  |  |  |
| None/some/neutral | 51.2 (950) | 43.1 (404) | 59.5 (546) |
| (very) large part | 48.8 (906) | 56.9 (534) | 40.5 (372) |
| **Connectedness to gay/queer-community** |  |  |  |
| Not connected/neutral | 43.0 (798) | 38.1 (357) | 48.0 (441) |
| Connected | 57.0 (1,058) | 61.9 (586) | 52.0 (477) |
| **<u>Behaviour</u>** |  |  |  |
| **Close physical contact with others in work or sports** |  |  |  |
| No | 76.6 (1,422) | 75.5 (708) | 77.8 (714) |
| Yes | 23.4 (434) | 24.5 (230) | 22.1 (204) |
| **Sex with men and women** |  |  |  |
| Only with men | 95.7 (1,777) | 97.1 (911) | 94.4 (866) |
| (also with) women in past 3 months | 4.3 (79) | 2.9 (27) | 5.7 (52) |
| **Number of male sex partners** past 3 months (median and IQR) | 3 [1-6] | 5 [3-10] | 2 [1-4] |
| **Anal sex with man without condom in past 3 months** |  |  |  |
| No | 34.5 (640) | 21.5 (202) | 47.7 (438) |
| Yes, with steady partner only | 22.4 (415) | 15.6 (146) | 29.3 (269) |
| Yes, (also) with casual sex partners | 43.2 (801) | 62.9 (590) | 23.0 (211) |
| **Practiced group sex in past 3 months with men** |  |  |  |
| No/not disclose | 73.0 (1,355) | 61.2 (574) | 85.1 (781) |
| Yes | 27.0 (501) | 38.8 (364) | 14.9 (137) |
| **Chemsex in past 3 months** |  |  |  |
| No/not disclose | 75.9 (1,409) | 64.4 (604) | 87.7 (805) |
| Yes | 24.1 (447) | 35.6 (334) | 12.3 (113) |
| **Received money or goods in exchange for sex past 3 months** |  |  |  |
| No/not disclose | 98.0 (1,818) | 97.0 (910) | 98.9 (908) |
| Yes | 2.0 (38) | 3.0 (28) | 1.1 (10) |
| **Sexual identity** |  |  |  |
| GBMSM | 98.7 (1,831) | 98.9 (928) | 98.4 (903) |
| TPG | 1.3 (25) | 1.1 (10) | 1.6 (15) |

& based on information in the questionnaire, which is a proxy for the Dutch MPX vaccination program criteria (at time of questionnaire 22 July-5 Sept 2022)

Of 938 PPV eligible respondents, 52% used HIV-PrEP and 24% were living with HIV (96% used ART) (Tab. 1). 28% knew someone who had MPX, 45% knew someone who was vaccinated against MPX, 38% lacked connectedness to the gay/queer-community, and over the past three months 40% reported group sex, 66% reported unprotected anal intercourse (UAI) during casual sex, and 70% reported more than three sex partners.

Of 918 PPV non-eligible respondents (HIV negative/untested; none used HIV-PrEP), 82% not attended a CSH in the past year, 48% lacked connectedness to the gay/queer-community, and over the past three months 15% reported group sex, 23% reported UAI during casual sex, 25% reported more than three sex partners (none engaged in SHC care), and 12% used drugs during sex.

### Willingness to accept vaccination

Of respondents, 81.5% were willing to accept vaccination; this was 85% in those PPV eligible and 78% in those non-eligible (Tab. 2). Of respondents, 12% were unwilling to accept vaccination; this was 10% in those eligible and 13.5% in those non-eligible. The remaining respondents (7%; 5%, 9%) were neutral. Of respondents, 90% were willing to accept PEPV; this was 90% in those PPV eligible and 90% in those non-eligible (Tab. 2).

**Table 2.** Proportion of respondents who reported (certainly or likely) willing to accept vaccination when offered, those who were neutral, and those who were reporting (likely or certainly) not willing to accept vaccination after being invited, by population subgroups, GBMSM/TGP participating in Dutch online MPX-survey

|  | Entire study population N=1,856 |  |  | Eligible for vaccination& (N=938) |  |  | Non-eligible for vaccination& (N=918) |  |  |
| --- | --- | --- | --- | --- | --- | --- | --- | --- | --- |
|  | willing to accept<br>% (n) | neutral<br>% (n) | unwilling<br>% (n) | willing to accept<br>% (n) | neutral<br>% (n) | unwilling<br>% (n) | willing to accept<br>% (n) | neutral<br>% (n) | unwilling<br>% (n) |
| **Willing to accept PPV** | 81.5% (1512)<br>(1,132 certainly, 380 likely) | 6.9% (128) | 11.6% (216)<br>(70 certainly, 146 likely) | 85.2% (799)<br>(666 certainly, 133 likely) | 5.0% (47) | 9.8% (92)<br>(30 certainly, 62 likely) | 77.7% (713)<br>(466 certainly, 247 likely) | 8.8% (81) | 13.5% (124)<br>(40 certainly, 184 likely) |
| **Willing to accept PEPV** | 89.8% (1667) | 4.5% (83) | 5.7% (106) | 90.0% (844) | 4.2% (39) | 5.9% (55) | 89.7% (823) | 4.8% (44) | 5.6% (51) |
| **By characteristics, Suppl. 2** |  |  |  |  |  |  |  |  |  |
| **<u>Sociodemographics</u>** |  |  |  |  |  |  |  |  |  |
| **Country of birth** |  |  |  |  |  |  |  |  |  |
| The Netherlands | 81.6 (1483) | 7.5 (128) | 12.6 (216) | 83.8 (648) | 5.7 (441) | 10.5 (81) | 75.9 (600) | 9.4 (74) | 14.7 (116) |
| Other | 90.1 (264) | 3.4 (100) | 6.5 (19) | 91.5 (151) | 1.8 (3) | 6.7 (11) | 88.3 (113) | 5.5 (7) | 6.3 (8) |
| **Residence country** |  |  |  |  |  |  |  |  |  |
| The Netherlands | 81.6 (1512) | 6.9 (128) | 11.5 (216) | 85.4 (689) | 4.9 (45) | 9.7 (89) | 77.7 (698) | 8.9 (80) | 13.4 (120) |
| Other | 29 (74.4) | 7.7 (3) | 17.9 (7) | 73.7 (14) | 10.5 (2) | 15.8 (3) | 75.0 (15) | 5.0 (1) | 20.0 (4) |
| **Residence level of urbanization** |  |  |  |  |  |  |  |  |  |
| Rural areas | 68.4 (80) | 12.8 (15) | 18.8 (22) | 75.0 (42) | 8.9 (5) | 16.1 (9) | 62.3 (38) | 16.4 (10) | 21.3 (13) |
| Hardly urbanized areas | 70.8 (97) | 9.5 (13) | 19.7 (27) | 72.3 (47) | 7.7 (5) | 20.0 (13) | 69.4 (50) | 11.1 (8) | 19.4 (14) |
| Moderately urbanized areas | 79.5 (140) | 11.4 (20) | 9.1 (16) | 83.5 (66) | 10.1 (8) | 6.3 (5) | 76.3 (74) | 12.4 (12) | 11.3 (11) |
| Strongly urbanized areas | 79.9 (306) | 6.0 (23) | 14.1 (54) | 83.3 (155) | 5.9 (11) | 10.8 (20) | 76.6 (151) | 6.1 (12) | 17.3 (34) |
| Extremely urbanized areas | 86.6 (844) | 4.9 (48) | 8.5 (83) | 89.9 (463) | 2.7 (14) | 7.4 (38) | 82.8 (381) | 7.4 (34) | 9.8 (45) |
| Unknown/abroad | 66.2 (45) | 13.2 (9) | 20.6 (14) | 70.3 (26) | 10.8 (4) | 18.9 (7) | 61.3 (19) | 16.1 (5) | 22.6 (7) |
| **Residence SES score neighborhood** |  |  |  |  |  |  |  |  |  |
| Low | 79.2 (481) | 7.6 (46) | 13.2 (80) | 84.1 (259) | 5.2 (16) | 10.7 (33) | 74.2 (222) | 10.0 (28) | 10.7 (6) |
| Middle | 80.0 (475) | 7.6 (45) | 12.5 (74) | 83.9 (203) | 6.1 (17) | 10.0 (28) | 76.4 (240) | 8.9 (28) | 14.6 (46) |
| High | 87.3 (495) | 4.6 (26) | 8.1 (46) | 89.7 (271) | 3.0 (9) | 7.3 (22) | 84.5 (224) | 6.4 (17) | 9.1 (24) |
| Unknown | 69.3 (61) | 12.5 (11) | 18.2 (16) | 70.8 (34) | 10.4 (5) | 18.8 (9) | 67.5 (27) | 15.0 (6) | 17.5 (7) |
| **Personal received education** |  |  |  |  |  |  |  |  |  |
| Low | 75.8 (91) | 13.3 (16) | 10.8 (13) | 81.3 (52) | 7.8 (5) | 10.9 (7) | 69.6 (39) | 19.6 (11) | 10.7 (6) |
| Medium | 77.1 (270) | 8.6 (30) | 14.3 (50) | 76.8 (136) | 7.9 (14) | 15.3 (27) | 77.5 (134) | 9.2 (16) | 13.3 (23) |
| High | 83.0 (1151) | 5.9 (82) | 11.0 (153) | 87.7 (611) | 4.0 (28) | 8.3 (58) | 78.4 (540) | 7.8 (54) | 13.8 (95) |
| **Age (in years)** |  |  |  |  |  |  |  |  |  |
| 16-30 | 78.9 (322) | 8,8 (36) | 12.3 (50) | 81.9 (131) | 5.0 (8) | 13.1 (21) | 77.0 (191) | 11.3 (28) | 11.7 (29) |
| 30-45 | 80.0 (485) | 6.3 (38) | 13.7 (83) | 84.1 (275) | 5.2 (17) | 10.7 (35) | 75.3 (210) | 7.5 (21) | 17.2 (48) |
| 45-55 | 83.2 (313) | 5.1 (19) | 11.7 (44) | 86.6 (187) | 4.2 (9) | 9.3 (20) | 78.8 (126) | 6.3 (10) | 15.0 (24) |
| >55 | 84.1 (392) | 7.5 (35) | 8.4 (39) | 87.7 (206) | 5.5 (13) | 6.8 (16) | 80.5 (186) | 9.5 (22) | 10.0 (23) |
| **Medical factors** |  |  |  |  |  |  |  |  |  |
| **Used HIV-PrEP** |  |  |  |  |  |  |  |  |  |
| No | 80.7 (1,102) | 7.2 (98) | 12.2 (166) | 86.8 (389) | 3.8 (17) | 9.4 (42) |  |  |  |
| Yes | 83.7 (410) | 6.1 (30) | 10.2 (50) | 83.7 (410) | 6.1 (30) | 10.2 (50) |  |  |  |
| **HIV status** |  |  |  |  |  |  |  |  |  |
| Negative/not tested or non disclose | 80.8 (1,318) | 7.4 (120) | 11.8 (193) | 84.9 (605) | 5.5 (39) | 9.7 (69) |  |  |  |
| Positive | 86.2 (194) | 3.6 (8) | 10.2 (23) | 86.2 (194) | 3.6 (8) | 10.2 (23) |  |  |  |
| **STI in past year** |  |  |  |  |  |  |  |  |  |
| No/ don't know | 80.1 (1,208) | 7.4 (112) | 12.5 (189) | 84.1 (525) | 5.3 (33) | 10.6 (66) |  |  |  |
| Yes | 87.6 (304) | 4.6 (16) | 7.8 (27) | 87.3 (274) | 4.5 (14) | 8.3 (26) |  |  |  |
| **Visited public health CSH clinic in past year** |  |  |  |  |  |  |  |  |  |
| No | 79.6 (801) | 8.0 (80) | 12.4 (125) | 85.2 (218) | 4.3 (11) | 10.5 (27) | 77.7 (583) | 9.2 (69) | 13.1 (98) |
| Yes | 83.6 (711) | 5.6 (48) | 10.7 (91) | 85.2 (581) | 5.3 (36) | 9.5 (65) | 77.4 (130) | 7.1 (12) | 15.5 (26) |
| **Vaccinated small pox** |  |  |  |  |  |  |  |  |  |
| No/don't know | 81.8 (830) | 6.4 (65) | 11.8 (120) | 85.7 (433) | 4.8 (24) | 9.5 (48) | 77.8 (397) | 8.0 (41) | 14.1 (72) |
| Yes | 81.1 (682) | 7.5 (63) | 11.4 (96) | 84.5 (366) | 5.3 (23) | 10.2 (44) | 77.5 (316) | 9.8 (40) | 12.7 (52) |
| **Overall rated health** |  |  |  |  |  |  |  |  |  |
| Not good/neutral | 81.1 (257) | 7.3 (23) | 11.7 (37) | 80.8 (118) | 6.2 (9) | 13.0 (19) | 81.3 (139) | 8.2 (14) | 10.5 (18) |
| (very) good | 81.5 (1,255) | 6.8 (105) | 11.6 (179) | 86.0 (681) | 4.8 (38) | 9.2 (73) | 76.8 (574) | 9.0 (67) | 14.2 (106) |
| **Had monkeypox since May 2022** |  |  |  |  |  |  |  |  |  |
| No/don't know | 81.4 (1,471) | 7.0 (126) | 11.7 (211) | 85.4 (770) | 5.0 (45) | 9.6 (87) | 77.4 (704) | 8.9 (81) | 13.6 (124) |
| Yes | 84.4 (38) | 4.4 (2) | 11.1 (5) | 80.6 (30) | 5.6 (2) | 13.9 (5) | 100.0 (9) | 0.0 (0) | 0.0 (0) |
| **<u>Social environment</u>** |  |  |  |  |  |  |  |  |  |
| Know someone with monkeypox |  |  |  |  |  |  |  |  |  |
| No | 79.5 (1,198) | 7.8 (117) | 12.7 (191) | 82.6 (557) | 5.6 (38) | 11.7 (79) | 77.0 (641) | 9.5 (79) | 13.5 (112) |
| Yes | 89.7 (314) | 3.1 (11) | 7.1 (25) | 91.4 (242) | 3.4 (9) | 4.9 (13) | 83.7 (72) | 2.3 (2) | 14.0 (12) |
| **Know someone who had been vaccinated** |  |  |  |  |  |  |  |  |  |
| No | 76.0 (879) | 9.1 (105) | 15.0 (173) | 79.2 (408) | 6.8 (35) | 14.0 (72) | 73.4 (471) | 10.9 (70) | 15.7 (101) |
| Yes | 90.6 (633) | 3.3 (23) | 6.2 (43) | 92.4 (391) | 2.8 (12) | 4.7 (20) | 87.7 (242) | 4.0 (11) | 8.3 (23) |
| **Interpersonal trust (have many people I can trust)** |  |  |  |  |  |  |  |  |  |
| Not (completely) agree | 82.1 (483) | 7.8 (46) | 10.0 (59) | 83.9 (251) | 5.7 (17) | 10.4 (31) | 80.3 (232) | 10.0 (29) | 9.7 (28) |
| (Completely) agree | 81.2 (1,029) | 6.5 (82) | 12.4 (157) | 85.8 (548) | 4.7 (30) | 9.5 (61) | 76.5 (481) | 8.3 (52) | 15.3 (96) |
| **Share of MSM in friend-social network** |  |  |  |  |  |  |  |  |  |
| None/some/neutral | 76.5 (727) | 9.2 (87) | 14.3 (950) | 80.2 (324) | 5.4 (22) | 14.4 (58) | 73.8 (403) | 11.9 (65) | 14.3 (78) |
| (very) large part | 86.6 (785) | 4.5 (41) | 8.8 (80) | 89.0 (475) | 4.7 (25) | 6.4 (34) | 83.3 (310) | 4.3 (16) | 12.4 (46) |
| **Connectedness to gay/queer-community** |  |  |  |  |  |  |  |  |  |
| Not connected/neutral | 74.4 (594) | 9.1 (73) | 16.4 (131) | 78.4 (280) | 7.3 (26) | 14.3 (51) | 71.2 (314) | 10.7 (47) | 18.1 (80) |
| Connected | 86.8 (918) | 5.2 (55) | 8.0 (85) | 89.3 (519) | 3.6 (21) | 7.1 (41) | 83.6 (399) | 7.1 (34) | 9.2 (44) |
| **<u>Behaviour</u>** |  |  |  |  |  |  |  |  |  |
| **Close non-sex physical contact with others** |  |  |  |  |  |  |  |  |  |
| No | 81.1 (1,153) | 6.8 (97) | 12.1 (172) | 84.9 (601) | 4.5 (32) | 10.6 (75) | 77.3 (552) | 9.1 (65) | 13.6 (97) |
| Yes | 82.7 (359) | 7.1 (31) | 10.1 (44) | 86.1 (198) | 6.5 (15) | 7.4 (17) | 78.9 (161) | 7.8 (16) | 13.2 (27) |
| **Sex with men and women** |  |  |  |  |  |  |  |  |  |
| Only with men | 81.6 (1,450) | 7.0 (124) | 11.4 (203) | 85.2 (799) | 4.9 (47) | 9.9 (90) | 77.8 (674) | 9.1 (79) | 13.0 (113) |
| (also with) women in past 3 months | 78.5 (62) | 5.1 (4) | 16.5 (13) | 85.2 (23) | 7.4 (2) | 7.4 (2) | 75.0 (39) | 3.8 (2) | 21.2 (11) |
| **Number of male partners past 3 months** |  |  |  |  |  |  |  |  |  |
| 0-1 | 67.5 (328) | 10.1 (49) | 22.4 (109) | 73.2 (82) | 4.5 (5) | 22.3 (25) | 65.8 (246) | 11.8 (44) | 22.5 (84) |
| 2-3 | 84.6 (411) | 7.0 (34) | 8.4 (41) | 85.0 (147) | 6.4 (11) | 8.7 (15) | 84.3 (264) | 7.3 (23) | 8.3 (26) |
| 4-5 | 88.4 (296) | 6.6 (22) | 5.1 (17) | 86.9 (193) | 6.3 (14) | 6.8 (15) | 91.2 (103) | 7.1 (8) | 1.8 (2) |
| >5 | 86.9 (477) | 4.2 (23) | 8.9 (49) | 87.5 (377) | 3.9 (17) | 8.6 (37) | 84.7 (100) | 5.1 (6) | 10.2 (12) |
| **Unprotected anal intercourse** |  |  |  |  |  |  |  |  |  |
| No | 80.0 (512) | 7.7 (49) | 12.3 (79) | 83.7 (169) | 5.4 (11) | 10.9 (22) | 78.3 (343) | 8.7 (38) | 13.0 (57) |
| Yes, with steady partner only | 74.7 (310) | 8.9 (37) | 16.4 (68) | 80.8 (118) | 6.8 (10) | 12.3 (18) | 71.4 (192) | 10.0 (27) | 18.6 (50) |
| Yes, (also) with casual sex partners | 86.1 (690) | 5.2 (42) | 8.6 (69) | 86.8 (512) | 4.4 (26) | 8.8 (52) | 84.4 (178) | 7.6 (16) | 8.1 (17) |
| **Practiced group sex past 3 months** |  |  |  |  |  |  |  |  |  |
| No/not disclose | 78.3 (1061) | 8.1 (110) | 13.6 (184) | 81.5 (468) | 6.1 (35) | 12.4 (71) | 75.9 (593) | 9.6 (75) | 14.5 (113) |
| Yes | 90.0 (451) | 3.6 (18) | 6.4 (32) | 90.9 (331) | 3.3 (11) | 5.8 (21) | 87.6 (120) | 4.4 (6) | 8.0 (11) |
| **Chemsex in past 3 months** |  |  |  |  |  |  |  |  |  |
| No/not disclose | 80.8 (1139) | 6.8 (96) | 12.3 (174) | 84.6 (511) | 4.6 (28) | 10.8 (65) | 78.0 (628) | 8.4 (68) | 13.5 (109) |
| Yes | 83.4 (373) | 7.2 (32) | 9.4 (42) | 86.2 (288) | 5.7 (19) | 8.1 (27) | 75.2 (85) | 11.5 (13) | 13.3 (15) |
| **Received money/goods in exchange for sex** |  |  |  |  |  |  |  |  |  |
| No/not disclose | 81.5 (1482) | 6.8 (123) | 11.7 (213) | 85.5 (778) | 4.6 (42) | 9.9 (90) | 77.5 (704) | 8.9 (81) | 13.5 (123) |
| Yes | 78.9 (30) | 13.2 (5) | 7.9 (3) | 75.0 (21) | 17.9 (5) | 7.1 (2) | 90.0 (9) | 0.0 (0) | 10.0 (1) |
| **Sexual identity** |  |  |  |  |  |  |  |  |  |
| GBMSM | 81.6 (1494) | 6.8 (124) | 11.6 (213) | 85.2 (791) | 5.1 (47) | 9.7 (90) | 77.9 (703) | 8.5 (77) | 13.6 (123) |
| TGP | 72.0 (18) | 16.0 (4) | 12.0 (3) | 80.0 (8) | 0.0 (0) | 20.0 (2) | 66.7 (10) | 26.7 (4) | 6.7 (1) |

& based on information in the questionnaire, which is a proxy for the Dutch MPX vaccination program criteria (at time of questionnaire 22 July-5 Sept 2022)

**Table 3.**
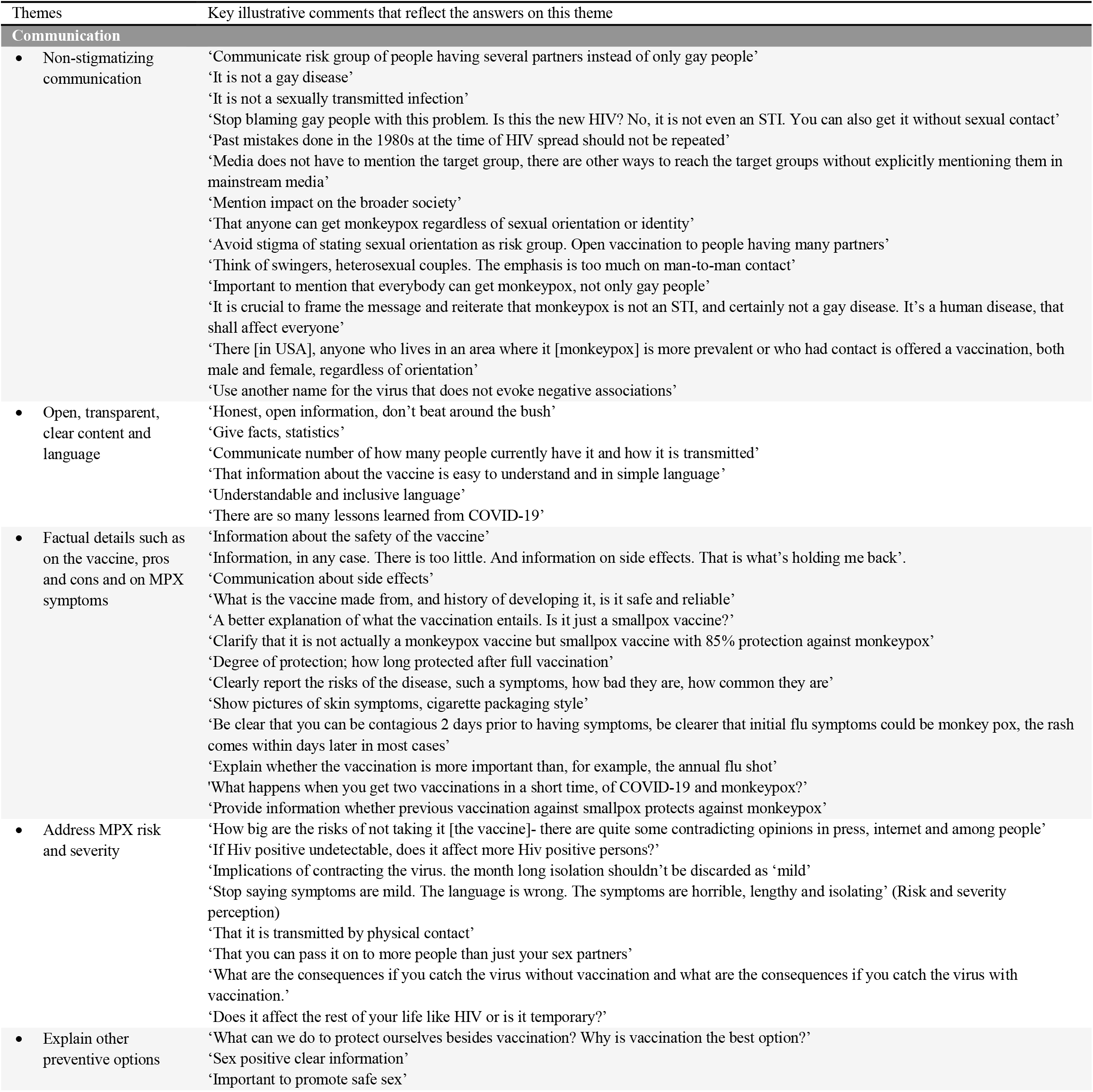
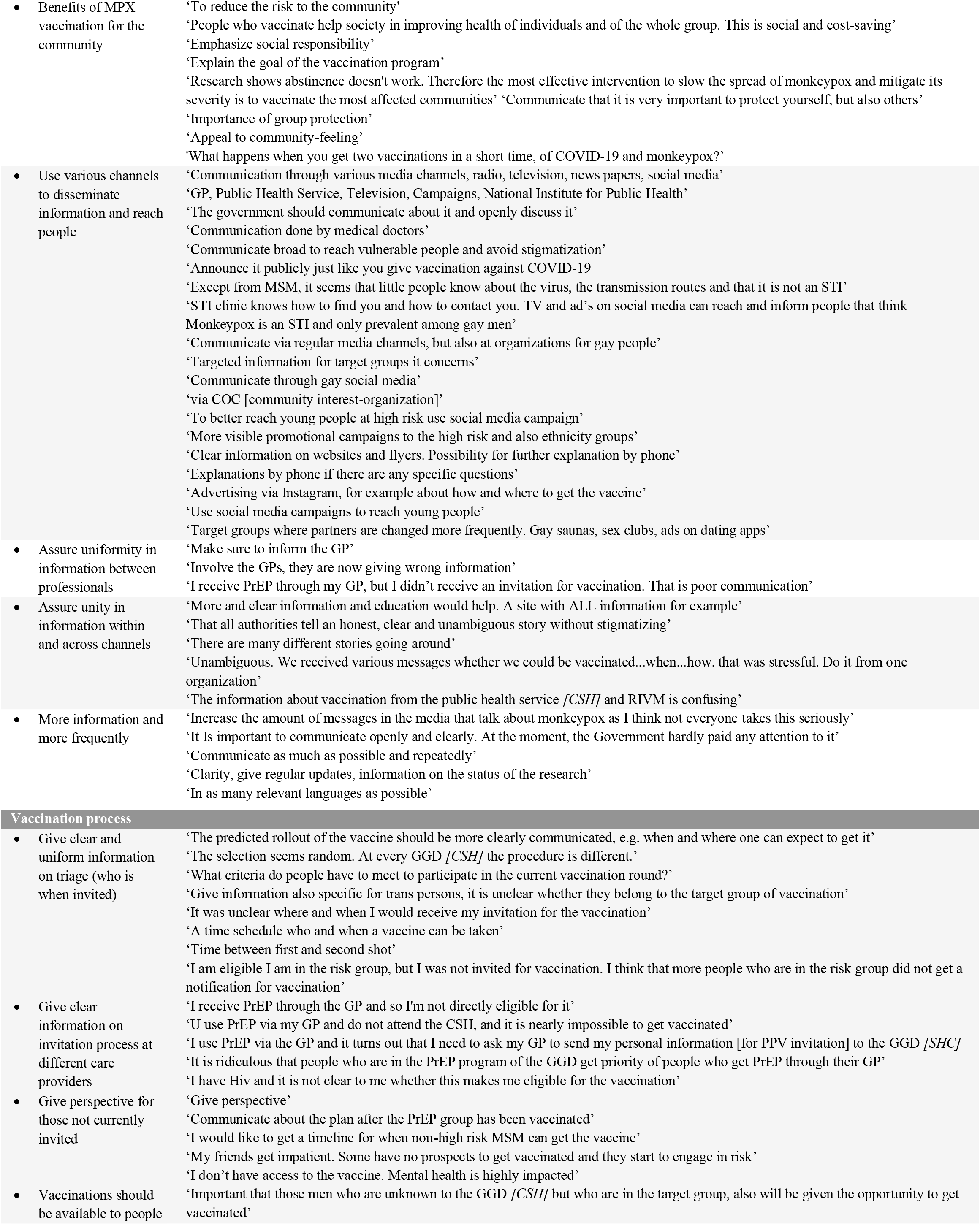
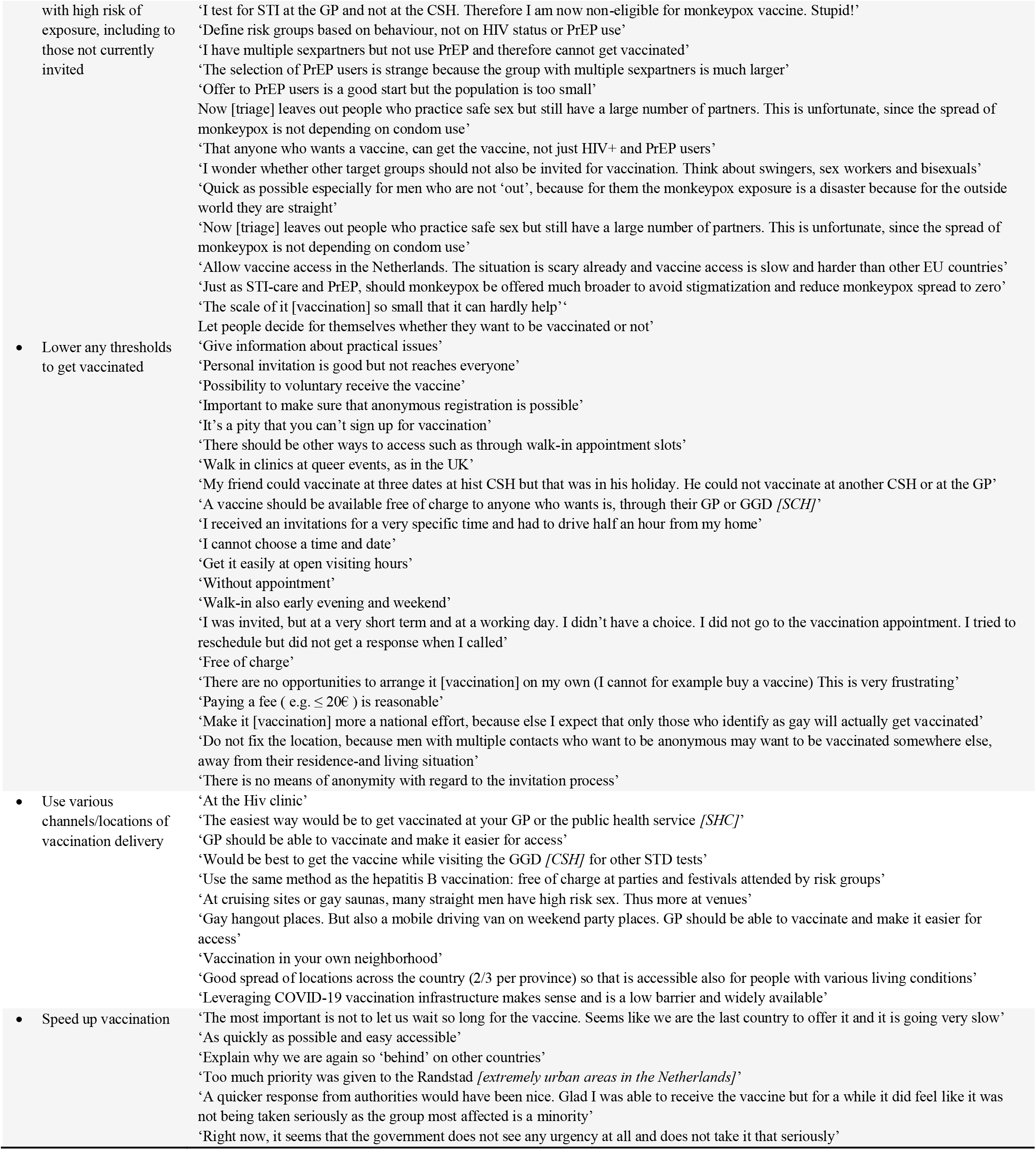
Recommendations for health-professionals and institutions to communicate on monkeypox vaccination and the monkeypox vaccination program, according to GBMSM/TGP participating in Dutch online MPX-survey

### Sociodemographic, medical, social, and behavioral determinants for vaccination non-acceptance

#### Univariable analyses

Proportions of respondents being willing to accept PPV, neutral, or unwilling were presented for each of the determinant-subgroups in Tab. 2.

The odds of being unwilling/neutral (versus being willing) were higher for those born in the Netherlands, who live in lower urbanized areas, or in lower socioeconomic status neighbourhoods, who have lower educational level (for ‘neutral’ only), or younger age (for ‘unwilling’ only), who are HIV negative/untested (for ‘neutral’ only), had past STI diagnosis, did not know a person with monkeypox or vaccinated for MPX, not had many MSM-friends in their friend-networks, lacked connectedness to the gay/queer community, had at most one recent sex partner, always used condoms/or practiced UAI only with a steady partner, or not recently had group sex.

These associated determinants were observed in the entire population; these were also observed in PPV-eligible respondents (except for neighbourhood socioeconomic status, age, HIV and STI status, and UAI) and in non-eligible respondents (except for educational level, age, and STI [HIV not evaluated]).

#### Multivariable analyses

The odds of being unwilling/neutral to accept PPV were higher for those born in the Netherlands, who live in lower urbanized areas, not knew MPX-vaccinated people, lacked connectedness to the gay/queer community, or who had at most one recent sex partner (Fig. 2). These determinants increased odds for both ‘unwilling’ and ‘neutral’ in the entire population and these same determinants were found associated in PPV-eligible and in non-eligible respondents (Fig. 2). For those eligible, being born in the Netherlands increased odds only for ‘neutral’, and having at most one partner increased odds only for ‘unwilling’. For those non-eligible, being born in the Netherlands, live in less urbanized areas [just borderline significant], or lack of connection to the gay/queer community only increased odds for ‘unwilling’.

**Figure 2.**
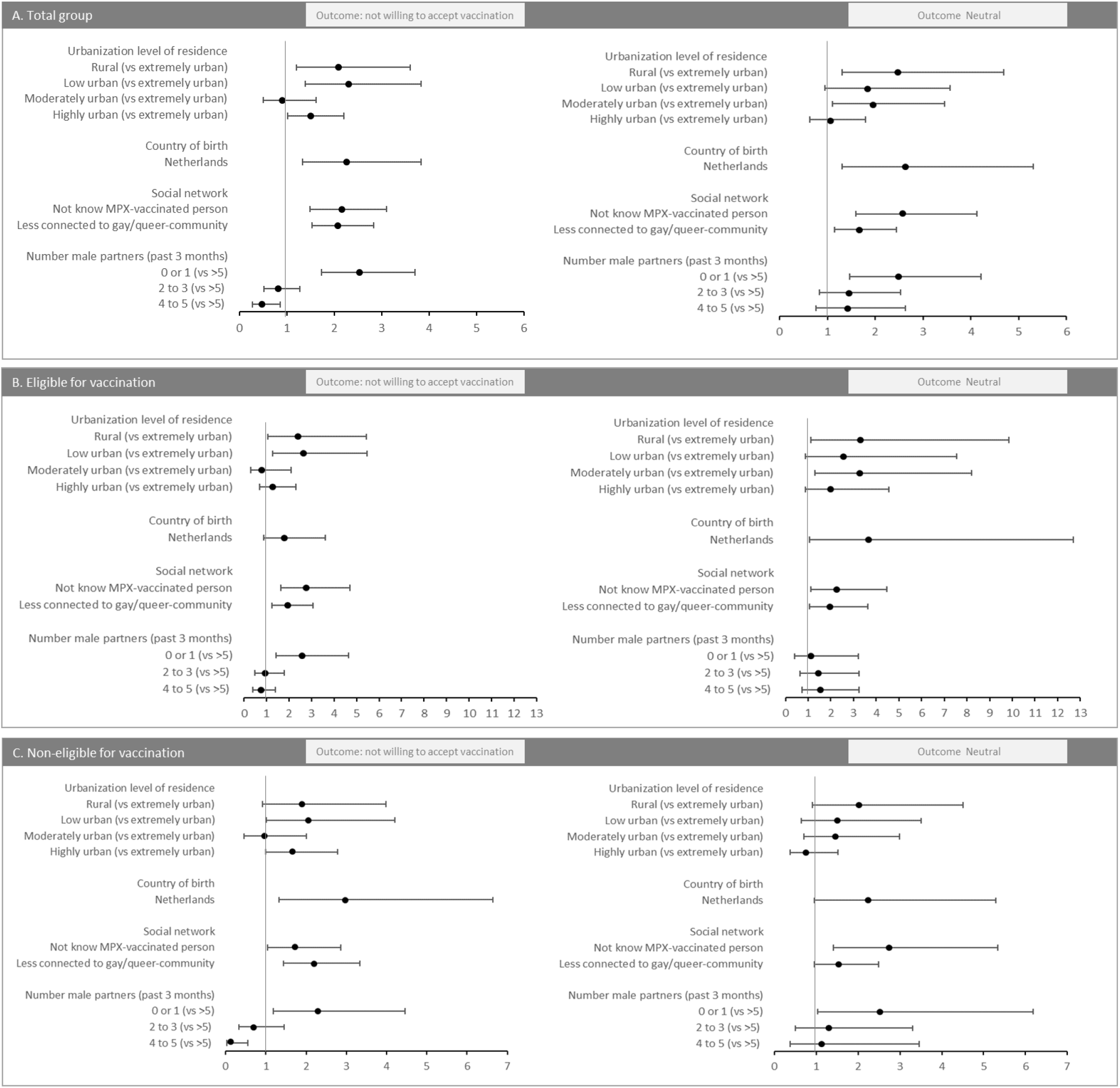
Sociodemographic, social environment, and behavioral determinants and their association with being not willing, or neutral, to accept monkeypox vaccination when offered, (compared to being willing), in multivariable multinominal regression analyses, MPX unvaccinated GBMSM/TGP participating in the Dutch online MPX-survey

**Figure 3.**
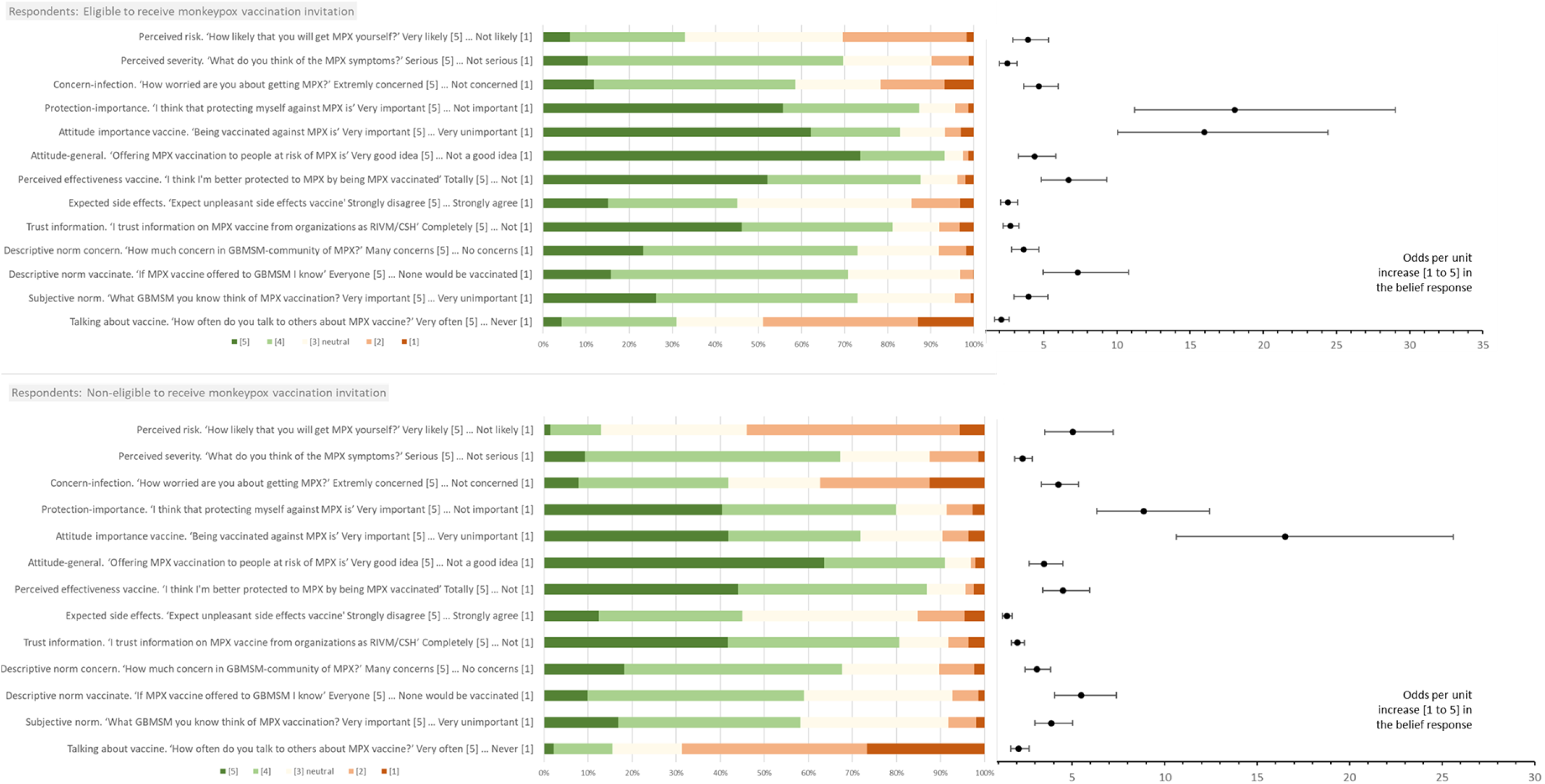
Distribution of responses to belief-statements in respondents who are eligible to receive PPV-invitation and those non-eligible (left panel), and the odds for each of these beliefs (right panel) for their association with being willing (certainly or likely) to accept monkeypox vaccination when offered (compared to not willing/being neutral), in multivariable logistic regression analyses, monkeypox unvaccinated GBMSM/TGP participating in Dutch online MPX-survey

### Beliefs and their association with willingness to accept vaccination

#### Beliefs distribution

Figure 2 presents the distribution of the response scores [1 to 5], for each belief. The proportion and number of respondents with score 4 or 5 were as follows:

Perceived risk, severity and concern about MPX: 32.9% (n=309) of eligible respondents and 13.0% (n=119) of non-eligible respondents considered themselves at risk for monkeypox, 69.7% and 67.2% thought that symptoms could be severe, and 58.5% and 41.8% were concerned about acquiring MPX.

Motivation/importance to protect against MPX: 87.3% of PPV eligible respondents and 79.8% of those non-eligible considered it important to protect themselves from MPX, 82.8% and 71.8% considered it important to be MPX vaccinated, 93.3% and 91.1% stated that vaccinating people with high risk was a good idea.

Perceived response efficacy of MPX vaccination: 87.6% of eligible and 86.9% of non-eligible respondents expected to be protected by the vaccine, 45.1% and 45.0% (score 1 or 2) did not expect severe side effects of vaccination, 81.1% and 80.6% had trust in information about MPX vaccine as provided by Dutch institutions.

Perceived social norms: 73.0% of eligible and 67.6% of non-eligible respondents thought the gay/queer community was concerned about MPX, 72.9% and 58.3% thought that many/most in their social network considered MPX vaccination important, 70.9% and 59.0% thought many/most in their social network would get MPX vaccinated, and 30.9% and 15.5% stated that they often discussed vaccination with others.

#### Association with willingness to accept vaccination

In both PPV eligible and non-eligible respondents, all evaluated beliefs were associated with being willing to accept PPV in both univariable and multivariable (Figure 2) logistic regression analyses. Multivariable models adjusted for country of birth, urbanization level, knowing MPX-vaccinated persons, connectedness to the gay/queer community, and number of sex partners.

### Community advice on communicating about monkeypox and PPV program

A total of 1,437 (52.8%) respondents filled in the open question to give their recommendations and two main themes arose from the answers, which were communication about MPX and the PPV program, and were related to (lifting) barriers regarding the access to information and to PPV.

#### Communication

Respondents recommended to provide information about MPX to increase public awareness/knowledge and to improve appraisal of personal risk and severity. Respondents mentioned the importance of communicating both pros and cons of MPX vaccination, degree of protection conferred, and the development of the vaccine (‘What is the vaccine made from, and history of developing it, is it safe and reliable’), and also how vaccination relates to other MPX prevention strategies (‘What can we do to protect ourselves besides vaccination? Why is vaccination the best option?’). Participants suggested to also explain other MPX preventive options but not advocating abstinence. Explain what is the goal of the PPV-program (note: public health goal of PPV is to limit MPX spread). Such communications could stimulate a positive attitude towards MPX vaccination and increase response efficacy of MPX vaccination, as well as protection by other preventive strategies.

Respondents stated the importance of non-stigmatizing language, such as linking infection risks to behaviors (such as multiple sexual partners), not to sexual orientation (‘it is not a gay disease’). It was found important to be more transparent and factual in communication (‘Honest, open information, don’t beat around the bush’; ‘Give facts, statistics’).

For optimal access to information for all people at high risk, respondents recommended to use mainstream media to underpin the importance of MPX and use public media to reach more ‘hidden’ target groups (‘Announce it publicly just like you give vaccination against COVID-19’), while others suggested community-specific channels (‘Communication through gay social media’). Respondents labelled discrepant information content across channels as disruptive, and they recommended more uniformity in communication-messages across the different communication channels, also have a central website, and with more frequent provision.

#### PPV-program

Respondents asked for more transparent information on who was eligible and when invited. Often respondents mentioned that the operational information around PPV was unclear and differed between healthcare providers(‘It was unclear where and when I would receive my invitation for the vaccination’). They also asked to communicate who is non-eligible and what is the prospect of receiving PPV later for the non-eligible person.

Participants stated that the clarity and uniformity in both the information and in the PPV procedures across healthcare settings and geographical regions could be improved (‘people who are in the PrEP program of the GGD [CSH] get priority over people who get PrEP through their GP’).

Respondents commented to improve PPV access and self-efficacy, with low-threshold options to get vaccinated. They recommended ‘self-registration’ (‘It’s a pity that you can’t sign up for vaccination’) in addition to personal invitations. Respondents suggested to offer vaccination at various clinic-and non clinic settings, also outside the region of residence, including anonymous and discrete PPV options, in neighbourhoods where people live (reduce travel distance), and where people get together (‘Gay hangout places. But also a mobile driving van on weekend party places. GP should be better informed and able to vaccinate and make it easier for access’), to offer vaccination at Hiv clinic, or to offer vaccination as done ‘with COVID-19’, ‘at test streets’ or as is routinely done during a the CSG visit (‘as with Hepatitis B’).

Respondents expressed concern that people with high risk for exposure are currently not invited for vaccination. For more inclusive vaccination-access, respondents frequently recommended to make PPV available for a broader group of people who had high risk. They mentioned to include people who not disclosed risk behavior to a healthcare professional or those who not engaged in preventive healthcare, and to offer PPV based on people’s willingness, rather than on categorization of people into subgroups (‘That anyone who wants a vaccine, can get the vaccine, not just HIV+ and PrEP users’). Respondents recommended to speed-up vaccination.

Other quotes regarding MPX communication and MPX PPV program are shown in Table 4.

## DISCUSSION

This survey was conducted at the start of the national MPX PPV program in the Netherlands in a convenience sample of GBMSM/TGP. This research assessed willingness and associated determinants for PPV (non-)acceptance, and collated respondents’ recommendations on campaigning and PPV-procedures, to inform public health strategies.

### Willingness to accept vaccination

Willingness to accept vaccination in this sample was high with 81.5% (86% in PPV eligible respondents). Another Dutch survey recruiting early July 2022 had observed 70% PPV willingness [21], which was confirmed in the current study showing 73% in the early recruitment at the end of July (Suppl. 3). Current study showed 86% PPV willingness in the first half of August. It should be noted that there was no independent time trend. Rather, fluctuations in willingness to accept vaccination over time are known to occur in an unfolding epidemic, with changing vaccine availability, media coverage, and number of vaccinated people over time, as was demonstrated for COVID-19 [22].

### Strategies to increase access to information and prevention

In the evolving public health response, strategies can be further optimized to ensure broad access to information for all people, and low threshold access to vaccination for people with high risk for monkeypox.

#### Tailoring communication messages to beliefs

The beliefs found important in MPX PPV acceptance were in line with those important in COVID-19 vaccination acceptance and previous surveys on MPX PPV acceptance [15,21-23], and included perceived risk/severity, motivation to protect against infection, perceived response efficacy of PPV, and perceived social norms. Of PPV-eligible respondents a third felt at risk (another third not felt at risk), over half were concerned, and majority (70%) thought that MPX symptoms could be severe.

Majority of respondents (over 80%) were motivated to prevent MPX and also were positive about PPV, but varied in their expectancy of side effects. Public health communication messages should include factual information on MPX (exposure risk, transmission routes, symptoms) and on the vaccine (side-effects, degree of protection for oneself and the community, history of development). This should help a person who has high risk for exposure to feel at risk and appraise MPX as potentially serious and PPV as beneficial in balance with risks of vaccinating (possible side effects) and of not vaccinating (health-and social impact of having MPX for self and others).

People need to have access to relevant facts to be able to make an informed autonomous choice [22]. and communication-messages should include information on PPV-program affairs (‘who, where, and when’ to increase self-efficacy to get PPV). Communication may further address perceived social norms. A majority of respondents (71-73%) thought that the gay/queer community was concerned about MPX and that those people they knew deemed PPV as important and would accept PPV. It is notable that part of PPV-eligible respondents never (13%) or only sometimes (36%) discussed vaccination with others. Discussions with social network members can be helpful to encourage preventive behavior, but people might expect or experience difficulties in talking to network members about MPX prevention. Previous research on STI testing learned that people may anticipate negative reactions (public stigma) and shame (self-stigma) when discussing STI and testing, and they avoid stigma by choosing to disclose only to single/few trusted peers (selective disclosure) [24].Difficulties to discuss MPX vaccination might also arise in relation to unequal and unclear vaccine-access. It is important that information is transparent about triage. Further, public health MPX communication strategies might be designed to encourage a person to talk about MPX prevention with a close trusted person, and be designed to leverage possible difficulties to enable discussion of the topic in the wider community.

#### Increasing the access to information

Messages can be delivered with the personal PPV invitation/reminder, which is the moment when the actual choice for PPV is made. Other channels include social media or websites which offer further benefits such as ease of maximizing dose and frequency of information exposure, and attractive (visual) tailoring to address problematic beliefs, to support decision making, norm setting, and maintenance of helpful beliefs over time [22]. Most (81%) respondents in the current survey trusted information from the institutions. They recommended multiple channels to disseminate information, including mainstream media, general health websites, at clinics, at venues (where people get together), and using specific community-based channels. Respondents thereby urged to pay attention to uniformity in the information across the different channels. Information-diffusion could also be promoted by the community itself in peer-to-peer activities, although this may be less suited to reach disconnected people. Important to note is that any chosen channel will also reach persons who not have access to PPV but who might become PPV-eligible in the future and who also could be social supporters to eligible persons in their PPV choice-process.

#### Lifting barriers to vaccination

To lift possible (regional-specific) barriers to getting vaccinated in PPV-invited people, respondents suggested to bring vaccination facilities close to a person’s home, at venues where people get together, offer discrete (anonymous) vaccination, provide vaccination at the sexual health check-up, and be able to actively self-register for getting a vaccination, in addition to a personal invitation for PPV.

#### Targeting subgroups

Subgroups less likely to want vaccination were defined by were they were born and live (born in the Netherlands, live in lower urbanized areas), their social networks (no MPX-vaccinated social network members, lack of connection to the gay/queer community), and by their sexual behavior (at most one recent partner). Disparities by urbanization level in MPX prevention behavior (PPV uptake and other MPX mitigation strategies) were demonstrated in two US studies [25,26], and was also observed in HIV testing uptake [27]. Public health efforts should be strengthened in less urbanized areas together with local stakeholder networks, community based organizations and local communities. Furthermore, respondents who lacked connectedness to the gay/queer community were less likely to want PPV, in line research findings on HIV testing and COVID-19 testing uptake [27,28]. Disconnected people are also known to less engage in preventive healthcare. Social connectedness is a strong factor in health and driver of prevention, as people may be supported by their peers and peers can be behavioral role-models [29-31]. Additional outreach efforts are needed to inform people who have a high risk of exposure and lack connection to the GBMSM/TGP community or to care, and who (according survey-respondents) may include bisexual men, sexworkers, migrant people, very young GBMSM/TGP, and those who not disclose as GBMSM/TGP including male swingers.

### Ensure access to information and prevention for people non-eligible for vaccination

People who currently not receive an MPX PPV invitation clearly have unmet needs regarding the prevention of MPX. Most of them did not engage in CSG/hospital care (GP was not asked for) and some reported recent group sex (15%), UAI in casual sex (23%), more than three sex partners (25%), or chemsex (12%). In the Netherlands, aim is to provide access to PPV for people at highest risk for monkeypox, these mainly include people with multiple sexpartners. In practice PPV access is organized by personal invitation based on information as available in existing patient registries [11]. While this allows to reach people in a relatively quick and feasible way, such strategy excludes those people at risk who not engaged in healthcare or had missing registry information.

Survey-respondents asked for a more inclusive PPV-access. In a US study among persons who not received MPX vaccination, more than a quarter had tried to get vaccinated [26]. In current study, vaccine-non-eligible respondents showed concern about acquiring MPX (42%), wanted to protect themselves against MPX (80%), and would accept PPV when offered (78%). It is important that low threshold vaccination strategies are explored, which allow access to PPV also for people who have high risk but who are not in existing invitation-selections.

In the challenging context of limited vaccine supply, communication-strategies have a crucial role in ensuring an equitable and inclusive access to information and access to preventive care options.

Expansion of the number of people that can receive PPV has been realized in the US and other countries by recent application of intradermal injection, after FDA and EMA has approved its use [32,33]. However, this will not be implemented in the Netherlands, as was stated in a recent policy brief [34]. Transparency about triaging is important [35].

As recommended by respondents, for all people who have a risk for exposure to MPX, regardless PPV-eligibility, information must be actively provided and easily accessible, with specific, non-stigmatizing and sex-positive guidance to enable people to act on the various MPX mitigation and care options (e.g., seek care for symptoms, and reduce close/intimate contact exposure risks) to prevent MPX acquisition, morbidity, and spread.

### Limitations

Limitations of this study should be recognized. Important subgroups, such as very young people, people with lower educational level, bisexual men, sexworkers, and TGP were underrepresented in this study, just as they are underrepresented in care. The sample is a convenience sample and not representative for all GBMSM/TGP in the Netherlands. Therefore, the main limitation is external validity, limiting generalizability of the overall proportion of willingness to vaccinate to the wider target population of GBMSM/TGP. We cannot rule out possible selection bias towards including respondents with a more positive attitude to PPV than the ‘general’ GBMSM/TGP population.

Strategies were taken to minimize selection bias and not to influence participation-interest or answers on beliefs, such as keeping communications in the survey text to strict a minimum. Further, selection bias might be introduced by differential drop-out of younger people who had fewer sexual partners.

That drop-outs were more often non-accepting of PPV was in line with the more frequent PPV non-acceptance observed in respondents with few sexual partners in the survey (Tab. 2). Furthermore, it should be noted that overall retention in this online survey was high with 89%.

An important general limitation hampering guidance on public health monkeypox preventive actions, is the lack of reliable (national, regional or subpopulation level) data on the number of people invited and vaccinated among invitees.

### Strengths

This study has several strengths. The study sample was substantial, and respondents had representation across a variety of subgroups and geographic areas. Another strength includes the timing of the survey and the assessment of a wide range of determinants at the early start of vaccination roll-out, which provided timely data to improve preventive strategies during the MPX vaccination program. A further asset is the theoretical underpinning of the research using a combination of theoretical behavioral change models to define determinants for PPV behavior.

Triangulation was applied of quantitative data on determinants and qualitative data on campaigning and procedural aspects of the PPV-program. Finally, a major strength of this study was the highly diverse and complementary composition of the research team, including scientists from fields of epidemiology, behavioral science, intervention design, implementation research, communication experts from public health centers and community based organizations, and healthcare professionals who serve the target population. In the context of a new epidemic and its public health response, this team-collaboration made it possible to quickly collect and process data, followed by immediate communication to policy makers and those involved in monkeypox prevention and PPV-program activities.

## Conclusion

In the current MPX epidemic, vaccinating those at high risk is a key public health measure. Efforts may be strengthened for those at risk but less likely to want vaccination, by regional approaches in less urbanized areas or outreach strategies for people who lack connection with the gay/queer community. Public health strategies will benefit from belief-tailored communication that is also transparent about PPV-triage and provides non-stigmatizing guidance for PPV and MPX mitigation strategies and healthcare options. In the context of limited vaccine supply in the Netherlands, public health strategies should be particularly careful to maintain equitable and inclusive access to broad preventive information and care options.

## Data Availability

The anonymous data for this study can be requested for research by sending an email to.

## Author Contributions

ND drafted the report and performed the statistical analyses and YE, FS, and ADA contributed to statistical analysis. ND, YE, FS, CdH, AN, AM, UD, EH, FS contributed to design of the survey. ND and YE coded the ‘qualitative’ data [on communication recommendations]. All authors reviewed the results, provided guidance, and drafted, reviewed, and provided critical feedback on the report.

## Acknowledgments

We are grateful to the following people and organizations for involvement in the recruitment of study participants: DC klinieken Lairesse Amsterdam (Hans-Erik Nobel), MUMC+ Department of infectious diseases, CSH Amsterdam (Adriaan Tempert and Justin Luidens), CSH South Limburg (Angelique Lahaut, Rocxanne Theuerzeit, Marita Werner, Ronald van Hooren), CSH Northern Limburg, CSH Utrecht (Mark van den Elshout), CSH Rotterdam-Rijnmond (Masja van der Pas and Charlotte Lantinga), CSH Gelderland-Zuid (B. Pool), CSH Ijsselland (Janine van den Brink), COC Limburg (Manuel Spier), Noordwest Ziekenhuisgroep (Frieda von Truien), and sex on premises-venues Limburg. Further, we are in depth to facilitating online social media recruitment to STI AIDS Netherlands (Sjoerd Visser and Laurian Kuiper), and John de Wit from University Utrecht for fruitful discussions, Kevin Konings for providing assistance with the data analyses, and Rianne Wit and Lisanne Steijvers for visualization. We are indebted to our community-panel members for collaboration in designing this study. Finally, we thank all participants for contributing with their invaluable comments and responses.

## References

1. Thornhill, J. P., Barkati, S., Walmsley, S., Rockstroh, J., Antinori, A., Harrison, L. B., Palich, R., Nori, A., Reeves, I., Habibi, M. S., Apea, V., Boesecke, C., Vandekerckhove, L., Yakubovsky, M., Sendagorta, E., Blanco, J. L., Florence, E., Moschese, D., Maltez, F. M., Goorhuis, A., … SHARE-net Clinical Group (2022). Monkeypox Virus Infection in Humans across 16 Countries - April-June 2022. The New England journal of medicine, 387(8), 679–691. https://doi.org/10.1056/NEJMoa2207323

2. Vaughan, A. M., Cenciarelli, O., Colombe, S., Alves de Sousa, L., Fischer, N., Gossner, C. M., Pires, J., Scardina, G., Aspelund, G., Avercenko, M., Bengtsson, S., Blomquist, P., Caraglia, A., Chazelle, E., Cohen, O., Diaz, A., Dillon, C., Dontsenko, I., Kotkavaara, K., Fafangel, M., … Haussig, J. M. (2022). A large multi-country outbreak of monkeypox across 41 countries in the WHO European Region, 7 March to 23 August 2022. Euro surveillance : bulletin Europeen sur les maladies transmissibles = European communicable disease bulletin, 27(36), 2200620. https://doi.org/10.2807/1560-7917.ES.2022.27.36.2200620

3. European Centre for Disease Prevention and Control. Epidemiological data on the 2022 monkeypox outbreak. Retrieved from ‘https://www.ecdc.europa.eu/en/monkeypox-outbreak’ [Online Resource] (last accessed 26/09/2022)

4. Edouard Mathieu, Fiona Spooner, Saloni Dattani, Hannah Ritchie and Max Roser (2022) - “Monkeypox”. Published online at https://OurWorldInData.org. Retrieved from: ‘https://ourworldindata.org/monkeypox’ [Online Resource] (last accessed 26/09/2022)

5. World Health Organization. News. ‘https://www.who.int/europe/news/item/23-07-2022-who-director-general-declares-the-ongoing-monkeypox-outbreak-a-public-health-event-of-international-concern’ [Online Resource] (last accessed 26/09/2022)

6. Hammarlund, E., Lewis, M. W., Carter, S. V., Amanna, I., Hansen, S. G., Strelow, L. I., Wong, S. W., Yoshihara, P., Hanifin, J. M., & Slifka, M. K. (2005). Multiple diagnostic techniques identify previously vaccinated individuals with protective immunity against monkeypox. Nature medicine, 11(9), 1005–1011. https://doi.org/10.1038/nm1273

7. Poland, G. A., Kennedy, R. B., & Tosh, P. K. (2022). Prevention of monkeypox with vaccines: a rapid review. The Lancet. Infectious diseases, S1473-3099(22)00574-6. Advance online publication. https://doi.org/10.1016/S1473-3099(22)00574-6

8. Petersen, E., Zumla, A., Hui, D. S., Blumberg, L., Valdoleiros, S. R., Amao, L., Ntoumi, F., Asogun, D., Simonsen, L., Haider, N., Traore, T., Kapata, N., Dar, O., Nachega, J., Abbara, A., Al Balushi, A., Kock, R., Maeurer, M., Lee, S. S., Lucey, D. R., … Koopmans, M. (2022). Vaccination for monkeypox prevention in persons with high-risk sexual behaviours to control on-going outbreak of monkeypox virus clade 3. International journal of infectious diseases : IJID : official publication of the International Society for Infectious Diseases, 122, 569–571. https://doi.org/10.1016/j.ijid.2022.06.047

9. World Health Organization. Vaccines and immunization for monkeypox: Interim guidance, 24 August 2022. Retrieved from ‘https://www.who.int/publications/i/item/WHO-MPX-Immunization-2022.2-eng’ [Online Resource] (last accessed 26/09/2022).

10. Nuzzo JB, Borio LL, Gostin LO. The WHO Declaration of Monkeypox as a Global Public Health Emergency. JAMA. 2022;328(7):615–617. doi:10.1001/jama.2022.12513

11. National Institute for public health and the environment Netherlands. Public information vaccination monkeypox [in Dutch] ‘https://www.rivm.nl/monkeypox-apenpokken/vaccinatie’ [Online Resource] (last accessed 26/09/2022)

12. Fernandez, M. E., Ruiter, R., Markham, C. M., & Kok, G. (2019). Intervention Mapping: Theory- and Evidence-Based Health Promotion Program Planning: Perspective and Examples. Frontiers in public health, 7, 209. https://doi.org/10.3389/fpubh.2019.00209

13. Kok, G (2014). A practical guide to effective behavior change: How to apply theory- and evidence-based behavior change methods in an intervention. Eur. Health Psychol. 2014;16:156–170. doi: 10.31234/osf.io/r78wh.

14. Peters G-J, Y (2014). A practical guide to effective behavior change: How to identify what to change in the first place. Eur. Health Psychol. 2014;16:142–155. doi: 10.31234/osf.io/hy7mj

15. Ten Hoor, G. A., Varol, T., Mesters, I., Schneider, F., Kok, G., & Ruiter, R. (2022). Just-in-Time, but Still Planned: Lessons Learned From Speeding up the Development and Implementation of an Intervention to Promote COVID-19 Vaccination in University Students. Health promotion practice, 15248399221095077. Advance online publication. https://doi.org/10.1177/15248399221095077

16. Rogers, R. W. (1975). “A protection motivation theory of fear appeals and attitude change”. Journal of Psychology. 91 (1): 93–114. doi:10.1080/00223980.1975.9915803. PMID 28136248.

17. Rogers, R.W. (1983). Cognitive and physiological processes in fear appeals and attitude change: A Revised theory of protection motivation. In J. Cacioppo & R. Petty (Eds.), Social Psychophysiology. New York: Guilford Press.

18. Fishbein, M. & Ajzen, I. (1975). Belief, attitude, intention, and behavior: An introduction to theory and research. Reading, MA: Addison-Wesley.

19. Ajzen, I (1991). The theory of planned behavior. Organizational Behavior and Human Decision Processes. 50 (2): 179–211. doi:10.1016/0749-5978(91)90020-T.

20. Luger, T.M. (2013). Health Beliefs/Health Belief Model. In: Gellman, M.D., Turner, J.R. (eds) Encyclopedia of Behavioral Medicine. Springer, New York, NY. https://doi.org/10.1007/978-1-4419-1005-9_1227

21. Wang, H., d’Abreu de Paulo, K., Gültzow, T., Zimmermann, H., & Jonas, K. J. (2022). Monkeypox self-diagnosis abilities, determinants of vaccination and self-isolation intention after diagnosis among MSM, the Netherlands, July 2022. Euro surveillance : bulletin Europeen sur les maladies transmissibles = European communicable disease bulletin, 27(33), 2200603. https://doi.org/10.2807/1560-7917.ES.2022.27.33.2200603

22. Sanders, J. G., Spruijt, P., van Dijk, M., Elberse, J., Lambooij, M. S., Kroese, F. M., & de Bruin, M. (2021). Understanding a national increase in COVID-19 vaccination intention, the Netherlands, November 2020-March 2021. Euro surveillance : bulletin Europeen sur les maladies ransmissibles = European communicable disease bulletin, 26(36), 2100792. https://doi.org/10.2807/1560-7917.ES.2021.26.36.2100792

23. Varol, T., Schneider, F., Mesters, I., Ruiter, R., Kok, G., & Ten Hoor, G. A. (2022). Facilitating Informed Decision Making: Determinants of University Students’COVID-19 Vaccine uptake. Vaccines, 10(5), 704. https://doi.org/10.3390/vaccines10050704

24. Theunissen, K. A., Bos, A. E., Hoebe, C. J., Kok, G., Vluggen, S., Crutzen, R., & Dukers-Muijrers, N. H. (2015). Chlamydia trachomatis testing among young people: what is the role of stigma?. BMC public health, 15, 651. https://doi.org/10.1186/s12889-015-2020-y

25. Hubach, R. D., & Owens, C. (2022). Findings on the Monkeypox Exposure Mitigation Strategies Employed by Men Who Have Sex with Men and Transgender Women in the United States. Archives of sexual behavior, 1–6. Advance online publication. https://doi.org/10.1007/s10508-022-02423-3

26. Delaney, K. P., Sanchez, T., Hannah, M., Edwards, O. W., Carpino, T., Agnew-Brune, C., Renfro, K., Kachur, R., Carnes, N., DiNenno, E. A., Lansky, A., Ethier, K., Sullivan, P., Baral, S., & Oster, A. M. (2022). Strategies Adopted by Gay, Bisexual, and Other Men Who Have Sex with Men to Prevent Monkeypox virus Transmission - United States, August 2022. MMWR. Morbidity and mortality weekly report, 71(35), 1126–1130. https://doi.org/10.15585/mmwr.mm7135e1

27. Leenen, J., Wijers, J., Den Daas, C., de Wit, J., Hoebe, C., & Dukers-Muijrers, N. (2022). HIV testing behaviour and associated factors in men who have sex with men by level of urbanisation: a cross-sectional study in the Netherlands. BMJ open, 12(1), e049175. https://doi.org/10.1136/bmjopen-2021-049175

28. Hammoud, M. A., Wells, N., Holt, M., Bavinton, B., Jin, F., Maher, L., Philpot, S., Haire, B., Degenhardt, L., Bourne, A., Saxton, P., Keen, P., Storer, D., & Prestage, G. (2022). COVID-19 Testing in a Weekly Cohort Study of Gay and Bisexual Men: The Impact of Health-Seeking Behaviors and Social Connection. AIDS and behavior, 1–9. Advance online publication. https://doi.org/10.1007/s10461-022-03831-1

29. Hawkinson, D. E., Operario, D., Hess, S., & van den Berg, J. J. (2022). Bridging the age gap: intergenerational communication of HIV risk and prevention among younger and older men who have sex with men. AIDS care, 1–7. Advance online publication. https://doi.org/10.1080/09540121.2022.2085865

30. Lyu, H., Zhou, Y., Dai, W., Zhen, S., Huang, S., Zhou, L., Huang, L., & Tang, W. (2021). Solidarity and HIV Testing Willingness During the COVID-19 Epidemic: A Study Among Men Who Have Sex With Men in China. Frontiers in public health, 9, 752965. https://doi.org/10.3389/fpubh.2021.752965

31. Valente, T.W. (2015). Social networks and health behavior. In: B. Rimer, K. Glanz, V. Vishwanath (Ed.). Health Behavior: Theory, Research & Practice. 6th Ed. Wiley; New York: 2015. pp. 205–222.

32. Larkin HD. FDA Authorizes Intradermal Vaccine, Streamlines Rules to Increase Monkeypox Treatment Access. JAMA. 2022;328(9):819. doi:10.1001/jama.2022.14692

33. European Medicine Agency. EMA’s Emergency Task Force advises on intradermal use of Imvanex / Jynneos against monkeypox. “https://www.ema.europa.eu/en/news/emas-emergency-task-force-advises-intradermal-use-imvanex-jynneos-against-monkeypox” [Online Content] (last accessed 26/09/2022)

34. Dutch government Policy brief ‘https://www.rijksoverheid.nl/binaries/rijksoverheid/documenten/kamerstukken/2022/09/29/kamerbrief-over-stand-van-zaken-apenpokken/kamerbrief-over-stand-van-zaken-apenpokken.pdf’ [Online document] (accessed 29-09-222)

35. Godlee F. Covid-19: Transparency and communication are key. BMJ. 2020;371:m4764. 10.1136/bmj.m4764

