## Supplement 1, 2 and 3 for "Monkeypox vaccination willingness, determinants, and communication needs in gay, bisexual, and other men who have sex with men, in the context of limited vaccine availability in the Netherlands (Dutch MPX-survey)"

### SUPPLEMENT 1
### RECRUITMENT CHANNELS

This study is a convenience sample of GBMSM and TGP, who were asked to fill in an online questionnaire. Recruitment was done 'offline' at clinics and sex-on-remises venues and online, via social media channels.

'Offline' venue-based

- Period: 22-07-2022 to 05-09-2022
- Contribution: Of the 1856 respondents included in the analyses 15.8% (n=294) were from this channel.
- How: Local venue-based. We provided recruitment materials (posters/pocket size cards with a QR-code to access the survey) at geographically spread Centers of Sexual Health clinics (of the Dutch Public Health Services), and HIV outpatient clinics (HIV-treatment centres) in Maastricht and Amsterdam. Community-organizations, such as the 'COC' Limburg also distributed the survey on their social networks. Also, recruitment materials were distributed at sex-on-premise venues such as saunas and parties in the region of Limburg between 15 July and 1 August. Furthermore, previous GBMSM respondents to a study on chemsex were approached by email to participate in the survey [Evers, Y. J., et al. (2019). PloS one, 14(5), e0216732. https://doi.org/10.1371/journal.pone.0216732]

Online social media-advertised

- Period: 01-08-2022 to 05-09-2022
- How: A digital recruitment strategy was developed in collaboration with experts in online research and digital marketers, including professionals from STI AIDS Netherlands, a national non-profit organization to promote sexual health information. The recruitment campaign consisted of banners ads displayed -per order of importance and volume - on a) social media (Facebook and Instagram) using stories and en linked ads displayed with priority to social media users interested in topics such as "gay love", well-known influencers and clothing-brands, and LGBTIQ+ areas of interest., b) gay dating sites or apps (Grindr and Hornet) through the sending of inbox messages with pictures of the campaign and textual explanation, c) on the sexual health promotion website www.mantotman.nl), and d) via the websites of community organizations (such as Hiv Vereniging, COC Nederland, Man tot Man, Bi+ Nederland, Club Church, Sauna Nieuwezijds, and others).
- Contribution: Of the 1856 respondents in the analyses 84.2% (n=1562) were from this channel.

**SUPPLEMENT 2**
**QUESTIONNAIRE: ORDER OF THE VARIABLES ASKED AND DETAILS ON MEASURES**

**The order of the questions in the online questionnaire.**
The questionnaire started with questions on sex, gender, and sex with man, for identifying GBMSM or other populations. All were asked to respond to the statement attitude-general. GBMSM then received the subsequent questions. These were the questions on previous recent MPX vaccination, and having had MPX. Then the statement of the main outcome followed, which was willingness to get vaccinated. After this statement, several belief-statements followed. Thereafter, the questions on sociodemographics, social environment, medical factors, and behavior were asked. Finally, postal code (4-digits) was asked to define level of urbanicity and socioeconomic status of the neighborhood. Finally, the questionnaire concluded with asking consent to be approached for follow-up study.

**The variables**
Sociodemographics.

- Age (in years category 16-30, 30-45, 45-55, and >55)
- Country of birth (Netherlands, other Western, non Western)
- Level of urbanization (based on 4-digit postal code: Rural (< 500 addresses per km2), Hardly urbanized (500 to 1000 addresses per km2), Moderately urbanized (1000-1500 addresses per km2), Strongly or extremely urbanized (1500-2500 addresses per km2), Extremely urbanized (>2500 addresses per km2), Unknown/abroad.
  Based on CBS: https://www.cbs.nl/en-gb/onze-diensten/methods/definitions/degree-of-urbanisation. den Dulk, CJ, H. van de Stadt, and JM Vliegen. 1992. "Een Nieuwe Maatstaf Voor Stedelijkheid: De Omgevingsadressendichtheid." Maandstatistiek van de Bevolking 40(7):14–27.
- Social economic score (SES) of residence (low, middle, high, based on tertiles, and unknown). Dutch SES scores based on income, education level and employment were extracted from the Netherlands Institute for Social Research (https://www.scp.nl) per four-digit postal code area of the respondent.
- Level of education (low, middle, high). Low: Primary education, prevocational secondary education (VMBO), the first three years of senior general secondary education (HAVO) or pre-university secondary education (VWO) and the secondary vocational assistant's training (MBO-1). Intermediate: Upper senior general or pre-university secondary education, basic vocational training (MBO-2), vocational training (MBO-3) and intermediate and specialist vocational training (MBO-4). High: University of applied sciences and research university education.
  Categorization according to CBS: https://www.cbs.nl/en-gb/news/2018/20/well-being-not-distributed-equally/education-level and https://www.cbs.nl/en-gb/news/2018/22/half-of-low-skilled-men-aged-25-44-are-smokers/level-of-educational-attainment

Behaviour.

- Having close (non-sex) physical contact with others in work or sports (no, yes)
- Sex with men only or also with women or with other gender identity in past 3 months
- Number of male sexpartners in past 3 months (regrouped into 0 or 1, 2 to 5, 6-10, >11, unknown)
- Anal sex with man without condom in past 3 months (no; yes, only steady partner; yes, (also) casual partner), groupsex in past 3 months (no/not disclose, yes)
- Chemsex in past 3 months [defined by use of cocaine, crystal meth, ghb/gbl, ketamine, speed, XTC, mdma, or designer drugs: 2-CB, 3MMC, 4-FA, 4-MEC] (no/not disclose, yes)
- Received money or goods in exchange for sex past 3 months (no/not disclose, yes).

Medical factors.

- Use of PrEP (no, yes in past 3 months, yes longer ago)
- HIV status (negative/untested/not disclose, positive [on ART or not on ART])
- STI history in past year, such as chlamydia, gonorrhea, or syphilis (no/unknown, yes)
- Visited STI clinic in past year (no, yes)

- Vaccinated for small pox (no/don't known, yes)
- Overall rated health (bad/neutral, good/very good)
- Had monkeypox since May 2022 (no, yes)
- Vaccinated for monkeypox (no, yes), date and reason

Social environment.

- Know someone who has (had) monkeypox (no, yes)
- Know someone who has been vaccinated against monkeypox (no, yes)
- Interpersonal trust: agree/completely agree with versus neutral/not agree/completely not agree to statement 'I have many people I can rely on completely'
- Share of MSM among own friends: everyone/a lot versus neutral/few/nobody to statement 'When you think about your friends, how many of them are MSM?' [nobody to everyone]
- Community-connectedness: (very connected/connected versus neutral/not connected/not connected at all to statement 'How connected do you feel to the gay/queer community?' [Not connected to very connected]

Willingness to accept vaccination

Intention to preventive vaccination (PPV) was operationalized as willingness to accept vaccination.
This was the main outcome in current study.
It was assessed using the statement 'If you could receive a vaccine against Monkeypox, would you get vaccinated against Monkeypox?' and measured by a 5-point Likert scale, with response options 'No, certainly not' [value 1]' 'No, probably not'[value 2], 'Neutral [value 3], 'Yes, probably [value 4], Yes, certainly [value 5]. Further, intention for post-exposure prophylaxis vaccination (PEPV) was assessed (same 5-point Likert scale) with the statement 'Suppose you had sex with someone with Monkeypox, would you get vaccinated?'.

Psychosocial determinants on beliefs.

These were also assessed using a 5-point Likert scale [1 to 5] by the following statements.

(i) Risk/severity perceptions and concern about MPX
- Perceived risk. 'How likely do you think it is that you will get infected with Monkeypox?' [Not likely at all to very likely]
- Perceived severity. 'What do you think of the Monkeypox symptoms?' [Not serious at all to very Serious]
- Concern-infection. 'How worried are you about getting Monkeypox?' [Not concerned at all to extremely concerned]

(ii) Motivation/importance to protect against MPX
- Protection-importance. 'I think that protecting myself against Monkeypox is' [Not important at all to very important]
- Attitude importance vaccine. 'Being vaccinated against Monkeypox is' [Very unimportant to very important]
- Attitude-general. 'I think that offering the Monkeypox vaccination to people at risk of Monkeypox is' [Not a good idea at all to a very good idea]

(iii) Perceived response efficacy of MPX vaccination
- Perceived effectiveness vaccine. 'I think I am better protected against contracting Monkeypox by being vaccinated with the Monkeypox vaccine' [Not at all to totally]
- [no] Expected side effects. 'I expect unpleasant side effects from the Monkeypox vaccine if I get vaccinated' [Strongly disagree to strongly agree] presented as reversed (because of hypothesized inverse association)
- Trust information. 'I trust the information about the Monkeypox vaccine from organizations such as National Institute for Public Health and the Environment (RIVM) and the Public Health Service (GGD)' [Not at all to completely]

(iv) Perceived social norms/influence

- Descriptive norm on concern. ‘How many concerns is there in the gay/queer community about the Monkeypox outbreak?’ [No concerns at all to many concerns]
- Subjective norm [attitude-importance]. ‘What do MSM you know think about vaccination about Monkeypox? They think that the vaccination is’ [Very unimportant to very important]
- Descriptive norm on getting vaccinated. ‘If vaccination against Monkeypox would be offered to MSM I know, I think that:’ [None would be vaccinated to everyone would be vaccinated]
- Talking about vaccine. ‘How often do you talk to others about the Monkeypox vaccine?’ [never to very often]

**SUPPLEMENT 3**
**SENSITIVITY ANALYSES OF RECRUITMENT FACTORS: TIME AND CHANNEL**

Recruitment spanned over a period from calendar week 29 (starting July 22 2022) through 35 (ending 5 September 2022).
In this survey period, different recruitment channels were applied in varying intensity over time, and also there were substantial societal changes related to the epidemic:

(1) Changes in the epidemiology: MPX cases substantially increased and then decreased again
(2) Changes in the availability of the MPX PPV: the first vaccinations were offered by CSH in extremely urban areas only, from 25 July onward. Regions in the rest of the country in the weeks thereafter started to invite people for vaccination, with variations in the timing and pace in which this happened.
(3) Changes in media coverage of the epidemic and the vaccination campaign. Communications varied over time to best address the target population and inform the about the situation, and also might have varied by regional CSHs.

Fluctuations in beliefs and intention over time is a common phenomenon in an unfolding epidemic, with changing vaccine availability, and media coverage over time, as was demonstrated for COVID-19 [Sanders, J. et al. (2021). Euro surveillance 26(36), 2100792. https://doi.org/10.2807/1560-7917.ES.2021.26.36.2100792].
In current survey, willingness to accept vaccination fluctuated between calendar weeks ranging between 72.6% (observed in week 29) and 90.3% (observed in week 31); it was 86.2% in the first half of August (week 30-32); Furthermore, it might be possible that different types of recruitment channels might attract respondents with different intention to vaccination. In current survey, willingness to accept vaccination was 73.8% (217/294) for offline recruitment and 82.9% (1295/1562) for online recruitment.
It is difficult to disentangle recruitment time and recruitment channel because actions on recruitment varied over time, and therefore we should be cautious to interpret the results in attempts to evaluate their effects. Nevertheless, to provide some insight we have performed a sensitivity analyses in which we checked in the multivariable models (that also included the key determinants, country of birth, urbanization, MPX-vaccinated people in the social network, connection to the gay/queer community, and number of partners), whether calendar week and channel type of recruitment were associated with being unwilling/neutral to accept vaccination. These analyses showed that these recruitment related factors were not independently associated (all $p>0.05$) in the entire population and eligibility groups. Adjustment of recruitment calendar weeks and channel did furthermore not substantially change the estimates observed for the other determinants in the multivariable models.
